# Study protocol: A national cross-sectional study on psychology and behaviour investigation of Chinese residents in 2024, PBICR

**DOI:** 10.1101/2025.01.14.25320520

**Authors:** Siyuan Fan, Yimiao Li, Fangjie Dong, Wenwen Chen, Qin Han, Tingyu Duan, Jinguang Zhang, Qiang Sun, Yanping Bao, Yujia Li, Shuang Zang, Xiaoming Zhou, Feng Jiang, Wai kit Ming, Xinying Sun, Yibo Wu

**Affiliations:** School of Public Health, Imperial College London, London, UK; School of Nursing, Tianjin Medical University, Tianjin, China; School of Journalism & Communication, Chongqing University, Chongqing, China; School of Politics and Public Administration, Wuhan University, Wuhan, China; College of Health Management, Shandong University of Traditional Chinese Medicine, Jinan, China; School of Journalism and Communication, Hebei Institute of Communications, Shijiwazhuang, China; Sun Yat-sen University, Guangzhou, China; Center for Health Management and Policy, School of Public Health, Shandong University, Shandong, China; National Health Development Research Center, Beijing, China; National Institute on Drug Dependence and Beijing Key Laboratory of Drug Dependence, Peking University, Beijing, China; School of Arts, Shandong University, Shandong, China; Department of Community Nursing, School of Nursing, China Medical University, Beijing, China; Shandong Provincial Hospital Affiliated to Shandong First Medical University, Jinan, China; Dongying People’s Hospital, Dongying, China; School of International and Public Affairs, Shanghai Jiao Tong University, Shanghai, China; Institute of Healthy Yangtze River Delta, Shanghai Jiao Tong University, Shanghai, China; Department of Infectious Diseases and Public Health, City University of Hong Kong, Hong Kong; School of Public Health, Peking University, Beijing, China

**Keywords:** Chinese residents, physical and psychological health, health behavior, cross-sectional study

## Abstract

**Introduction:** This study protocol specifies the primary research line and theoretical framework of the 2024 Survey of the Psychology and Behavior of the Chinese Population. It aims to establish a consistent database of Chinese residents’ psychological and behavioral surveys through multi center and large sample cross sectional surveys to provide robust data support for developing research in related fields. It will track the public’s physical and psychological health more comprehensively and systematically.

**Methods:** The study was conducted from June 23, 2024 to September 29, 2024, using stratified and quota sampling methods. A total of 150 cities across 800 communities/villages were surveyed, selected from China (Despite extensive coordination, we have been unable to contact our counterparts in the Taiwan region of China to obtain relevant statistical data). The questionnaires were distributed to the public one on one and face to face by trained surveyors. The questionnaires included basic information about the individual, personal health status, basic information about the family, the social environment in which the individual lives, psychological condition scales, behavioral level scales, other scales, and attitudes towards topical social issues. Supervisors conducted quality control during the distribution process and returned questionnaires, logically checked and cleaned for data analysis.

**Discussion:** Data collection has been finished, and scientific outputs based on this data will support the development of health promotion strategies in China and globally. In the aftermath of the pandemic, it will guide policymakers and healthcare organizations to improve their existing policies and services to maximize the physical and mental health of the Chinese population.

## 1 Introduction

A cross-national study mentioned that lifetime prevalence of any mental disorder was 28.6% for males and 29.8% for females which were not a low proportion, and showed the grim situation of mental health[1]. However, according to the World Health Organization (WHO), mental health is a lot more than the absence of illness: it is an intrinsic part of our individual and collective health and well-being. In recent years, people pay more and more attention on mental health, especially after the outbreak of COVID-19 in 2020, as they may experience loneliness, denial, anxiety, depression, insomnia, and despair consequently[2]. As estimated, it has put the rise in both anxiety and depressive disorders at more than 25% during the first year of the pandemic[3]. This significant increase underscores the critical importance of prioritizing mental health and implementing comprehensive support and intervention strategies to address the growing mental health crisis. Different definitions of health behavior exist globally, and the WHO emphasizes that human behavior has a substantial impact on health outcomes[4]. Health behaviors can be classified into two categories: positive and negative. These behaviors are actions that individuals take to maintain or improve their health. They play a significant role in shaping physical and mental well-being and can influence the risk of chronic diseases and overall quality of life. It contains but not limited to physical activity, dietary habit, social media addiction, smoking and so on[5].

Mental health and health behaviors are interdependent. Poor mental health can affect an individual’s behavioral decisions, while a lack of healthy behaviors can compromise the physiological basis of mental health. The interaction between these two factors significantly impacts overall health outcomes[6]. Therefore, the mental health issue should arouse the whole society’s attention. There’s various factors are able to influence it. For example, the study conducted by Diane C Lim, MD et al[7], illustrate that the disturbed sleep is associated with a broad set of health outcomes. Also, a longitudinal Studies of Young People in England, conducted by Tayla McCloud et al[8]. aimed to assess the association between mental health and education level and found that in young people who attended higher education at ages 18-20 years had higher symptoms of common mental disorders at ages 16-17 years than those who did not. The experiences of trauma in childhood also influence the mental disease[9]. Health behaviors are also influence by multiple elements. Katrina Wilhite et al[10]. conducted a systematic review and found that a high level of physical activity and a low level of sedentary behavior were associated with the best physical health, psychological health. However, in the previous study, both study of mental health and health behaviors focus on the specific areas or particular crowd, which highlights the present research’s limitations.

In summary, there is no extensive, nationwide survey that examines the psychological and behavioral health status of a large and diverse population. The PBICR examines various aspects of mental health and health behaviors in China, ensuring timeliness and efficiency. It promotes open data access and sharing, and builds a high-quality, large-sample, multi-center, repetitive, nationwide cross-sectional database[6]. The 2024 PBICR project includes significant and relevant variables in the field of psychiatry, such as anxiety, depression, self-injury, and suicide[11]. This initiative aims to provide robust data support for research and development in various fields, enable a more comprehensive and systematic understanding of the public’s physical and mental health, and inform policy-makers and healthcare organizations to improve public health initiatives and enhance the well-being of residents and their families.

## 2 Methods

### 2.1 Study Design and Setting

This cross-sectional survey was initiated by the School of Public Health of Peking University. A total of 150 cities, 202 districts/counties, 390 townships/towns/streets, and 800 communities were randomly selected from 4 municipalities directly under the Central Government, 22 provinces, 5 autonomous regions, and 2 special administrative regions (Hong Kong and Macao) from June 20 to September 29, 2023. At present, this study has been approved by the Key Laboratory of Health Economics and Policy Research of the National Health Commission (NHC-HEPR202401). It has passed the ethics review of Shanghai Jiao Tong University (H20240237I). The study has also been registered in the Chinese Clinical Trials Registry (ChiCTR).

In this investigation, the research team designed the questionnaire after scientific and comprehensive review of books and literature, conducted several network expert consultations and discussions before the formal use of the questionnaire, and conducted three rounds of pre-survey. The research team collected and sorted out the feedback opinions in time, and modified them after discussion. At the same time, the research group selected the members of the provincial volunteer team working committee after strict resume screening and online interview. After completing three rounds of training and passing the test, a formal investigation could be started.

### 2.2 Participants

Participants must have the nationality of the People’s Republic of China and be a permanent resident of China (annual departure practice ≤1 month). Participants must be at least 18 years of age and have the ability to understand the meaning of each item in the questionnaire, and can complete the online questionnaire on their own or with the help of the investigator. All participants participated in the study voluntarily and filled out the informed consent form. People who are delirious, mentally abnormal, and have cognitive disabilities will be excluded, as will those who are participating in other similar studies or do not want to participate in the study.

In order to ensure the quality control of every aspect of the project, we have set up a complete organizational framework to execute every aspect of the project. The organizational framework of this survey project follows a general manager system, wherein each team is led by an individual who oversees and coordinates their collaborative efforts to complete the survey project. The research team comprises five distinct research groups, namely the expert committee, survey group, training and coordination group, scale design group, and quality control group. This year, we established a dedicated Investigator working committee to better align with PBICR’s long-term construction plan and facilitate further development of the field; It is also conducive to efficient work and management, which can effectively improve quality control and ensure that our projects and research meet higher standards.

Investigators will take the initiative to contact local community health centers or neighborhood committees, establish investigation sites, and issue electronic questionnaires through one-on-one on-site interviews. During the survey, investigators recruited participants. They also verify that participants meet study inclusion criteria and do not meet exclusion criteria. The questionnaire information completed by participants will be automatically summarized to the background server. The e-questionnaire was produced through the online survey platform (https://www.wjx.cn/), and respondents answered by clicking on the link. For respondents with limited mobility, investigators will conduct one-on-one interviews and fill out questionnaires on their behalf.

### 2.3 Quality Control

In this study, quality control is mainly carried out in five stages: questionnaire design process, pre-investigation process, investigators’ training process, questionnaire distribution process and questionnaire retrieval and analysis process.

#### 2.3.1 Questionnaire design Process

The survey team designed the questionnaire after scientific and comprehensive review of books and literature, and conducted online expert consultation and discussion among 48 experts from March to June 2024 before the formal use of the questionnaire. The experts consulted were all with senior professional titles and regional and professional representatives. Professional scope includes social medicine, behavioral epidemiology, psychology, health education, health statistics, health management, humanities medicine, journalism and communication, clinical medicine, pharmacy, nursing, sociology, philosophy, etc. According to the expert opinions, adjust the feedback and issue three rounds of pre-questionnaire.

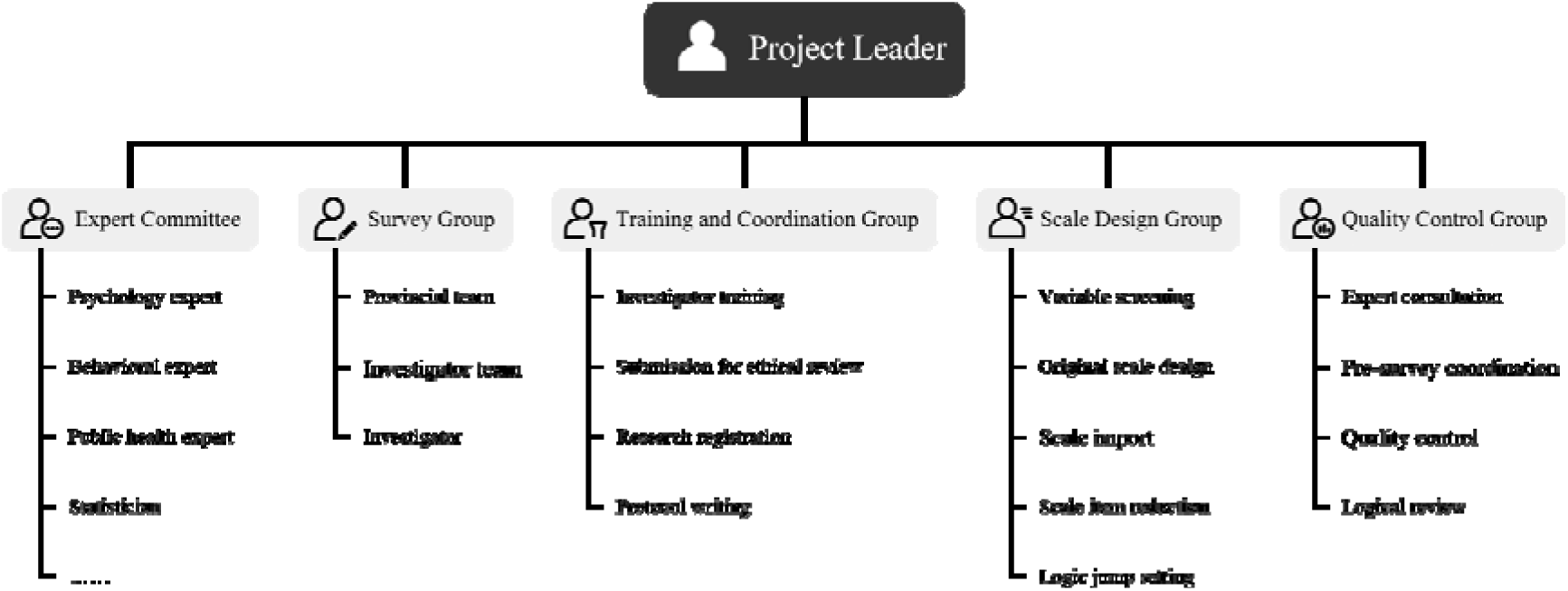

#### 2.3.2 Pre-investigation Process

This study conducted three rounds of pre-experiments from May 25 to May 28, May 30 to June 2, and June 4 to June 8, 2024, respectively. The sampling method of pre-survey was quota sampling, and the quota attributes were the same as the requirements of the formal survey, and the samples were 100, 100 and 200 people each time. During the pre-investigation period, feedback from respondents was collected and sorted out in time, and modified after discussion by members of the research group. The final questionnaire was revised after three rounds of preliminary survey. The questionnaires collected in the pre-survey stage were not included in the final analysis.

#### 2.3.3 Investigators’ Training Process

This study implements a “two-tier recruitment and hierarchical training” strategy to establish a high-quality survey team. Initially, provincial coordinators are selected through online recruitment, resume screening, and comprehensive interviews, evaluating their research competence, management capabilities, communication skills, decision-making abilities, stress resilience, and language adaptability. A minimum of two coordinators are assigned to each province, municipality, or autonomous region. Selected candidates undergo systematic training over several weeks, covering project overview, research objectives, survey methodology, questionnaire design, and other essential aspects, with a final assessment determining their qualification.

Subsequently, provincial coordinators are responsible for recruiting and training investigators within their respective regions. They develop training programs encompassing survey purposes and significance, questionnaire completion guidelines, field survey techniques, and data quality control measures. Additionally, they establish evaluation mechanisms to ensure that only qualified investigators who pass the assessment are authorized to conduct specific survey tasks.

#### 2.3.4 Questionnaire Distribution Process

To ensure the rigor of data collection, the research team has implemented multi-faceted quality control strategies. Initially, a strict questionnaire coding system was established, assigning unique identifiers traceable to respondents, investigators, and provincial coordinators. Questionnaires that fail to comply with the coding rules are deemed invalid to maintain dataset integrity and consistency. Furthermore, all investigators are required to distribute and collect questionnaires through face-to-face interactions to maximize validity. The research team has established a regular communication mechanism. Between July 4 and September 6, 2024, online meetings are conducted with provincial coordinators every Friday at 8 PM via the Tencent Meeting platform. These sessions involve reviewing and evaluating the weekly collected questionnaires, addressing emerging issues, responding to queries, and facilitating real-time discussions on challenging or controversial items. Additionally, the team implements multi-level quality control measures, including regular questionnaire reviews to ensure data accuracy and completeness. Professional data management software is utilized for secure storage and effective management, with regular backups to prevent data loss.

#### 2.3.5 Questionnaire Retrieval and Analysis Process

The research team has developed and implemented a rigorous questionnaire validation procedure, incorporating both quantitative and qualitative assessment methods. Each questionnaire undergoes independent logic verification by two professionally trained investigators to minimize human error and ensure assessment consistency. The validation criteria encompass several key dimensions: (1) Time validity check: Based on pretest results and expert evaluation, a minimum valid completion threshold of 300 seconds is established; (2) Internal consistency check: Identifying logical contradictions within questionnaires, such as consistency in demographic information and logical relationships between political affiliation and religious beliefs; (3) Completeness check: Questionnaires with a missing value ratio exceeding 20% are deemed informationally incomplete; (4) Duplication check: Advanced data analysis techniques are employed to cross-compare all collected questionnaires for identifying and eliminating duplicate submissions; (5) Response pattern analysis: Specialized algorithms are developed to detect abnormal response patterns, such as straight-line answers or other systematic patterns.

During the data analysis phase, the team employs advanced statistical methods for comprehensive analysis. For detected outliers or anomalous data points, the team traces back to original questionnaires or communicates directly with relevant investigators to verify information accuracy. Additionally, a dynamic questionnaire review mechanism is established, whereby new logical inconsistency patterns discovered during the survey process prompt immediate updates to screening criteria and retrospective reviews of collected questionnaires.

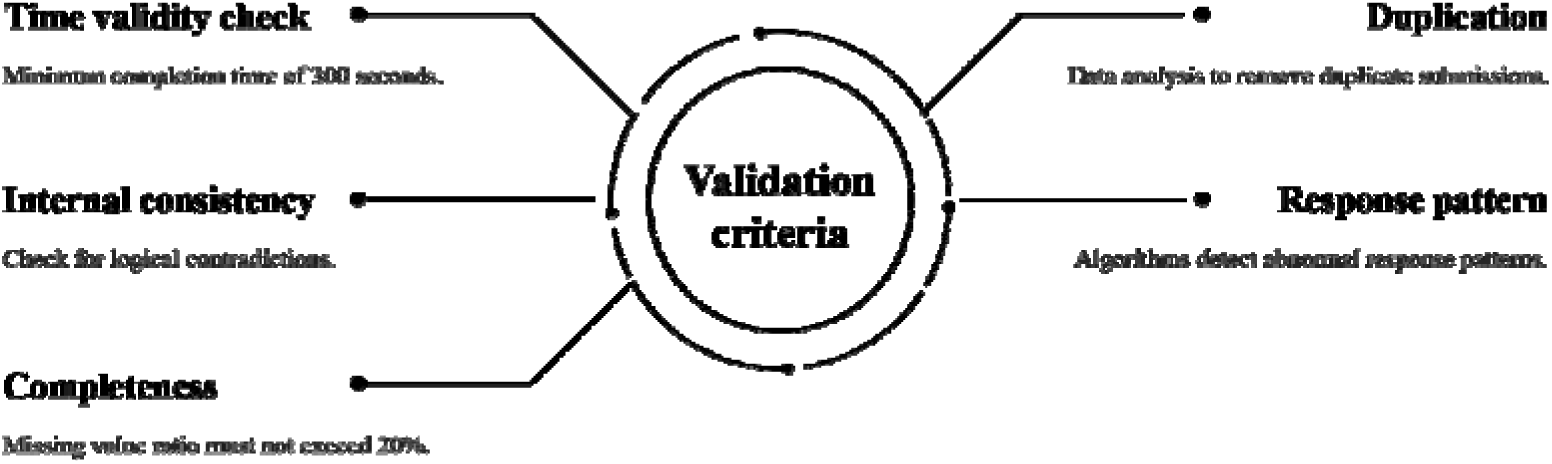

**Figure 1.**
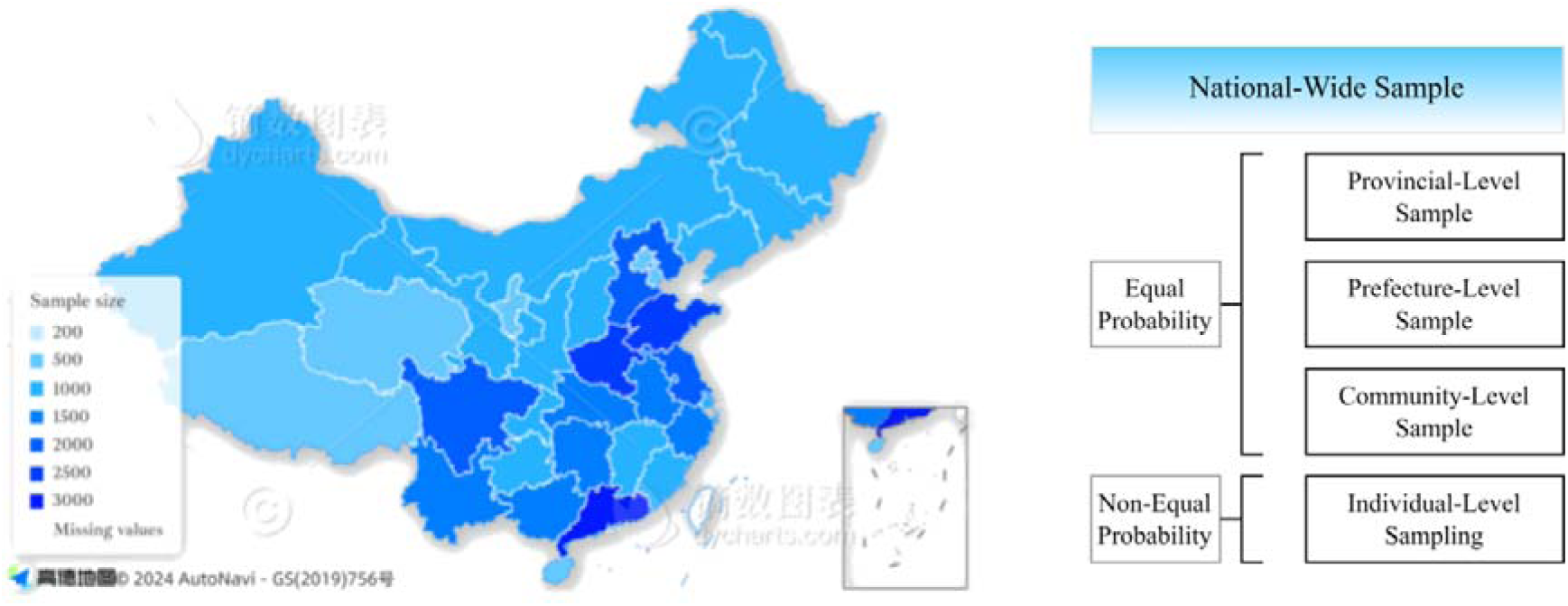
Sample screening process.

**Figure 2.**
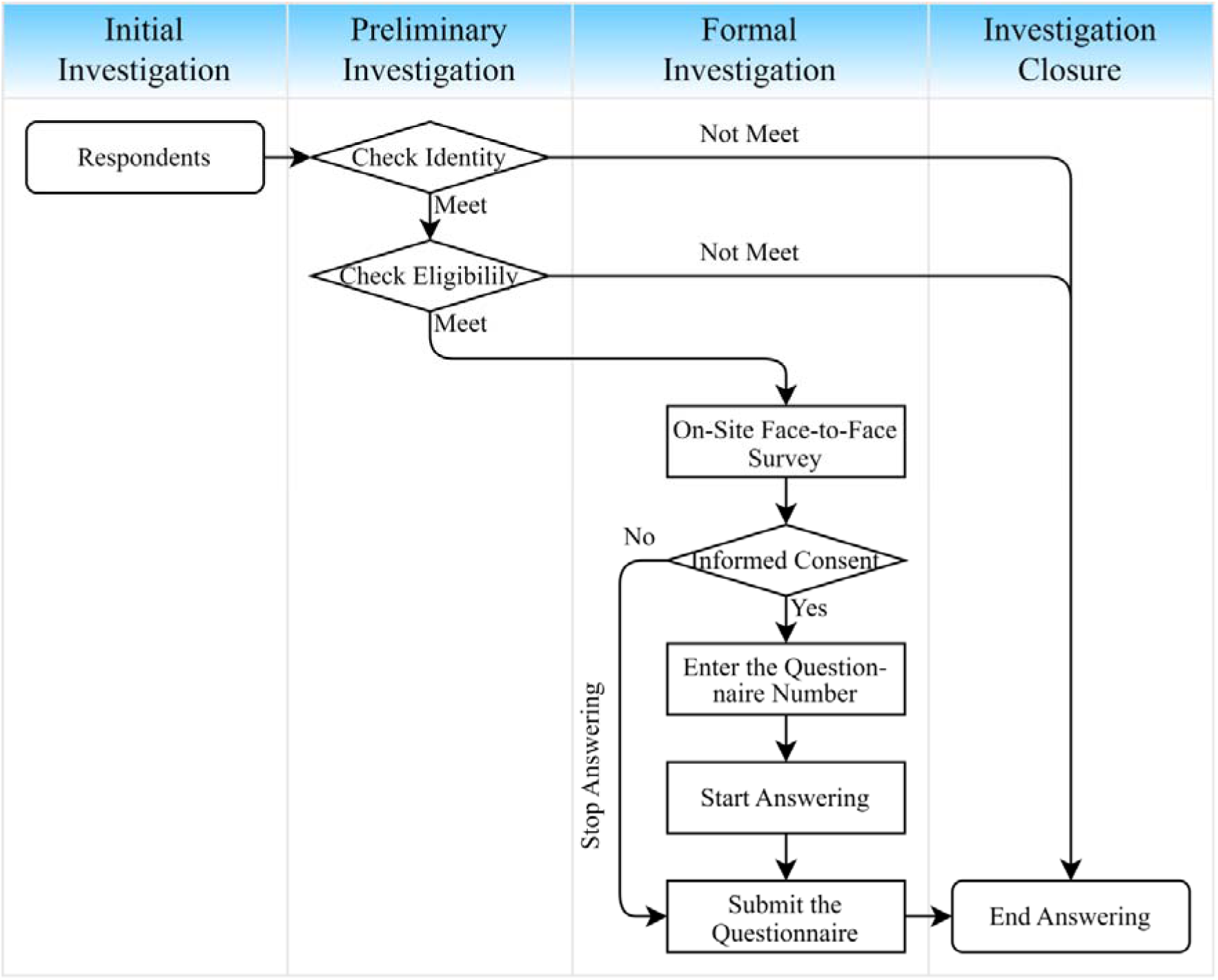
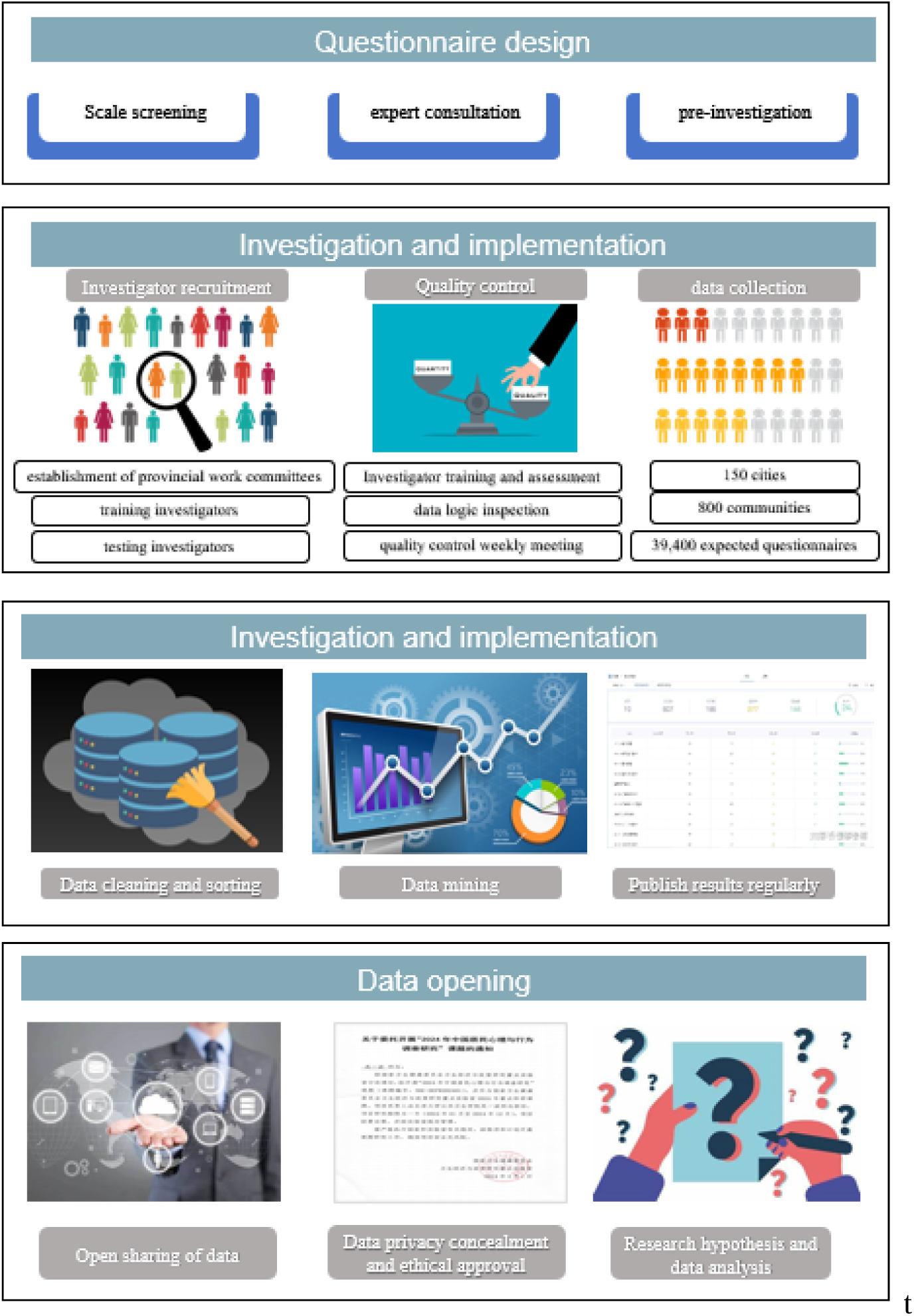
dInvestigation process.

### 2.4 Measurements

In addition to the informed consent form and questionnaire number, the questionnaire for this study comprised nine principal sections: basic personal information, personal health status, basic family information, social environment scale, psychological dimension scale, behavioural dimension scale, other scales, childhood experience questionnaire (self-administered), and attitudes towards topical social issues. Table 1 provides a detailed overview of the content and structure of each scale.

**Table 1.**
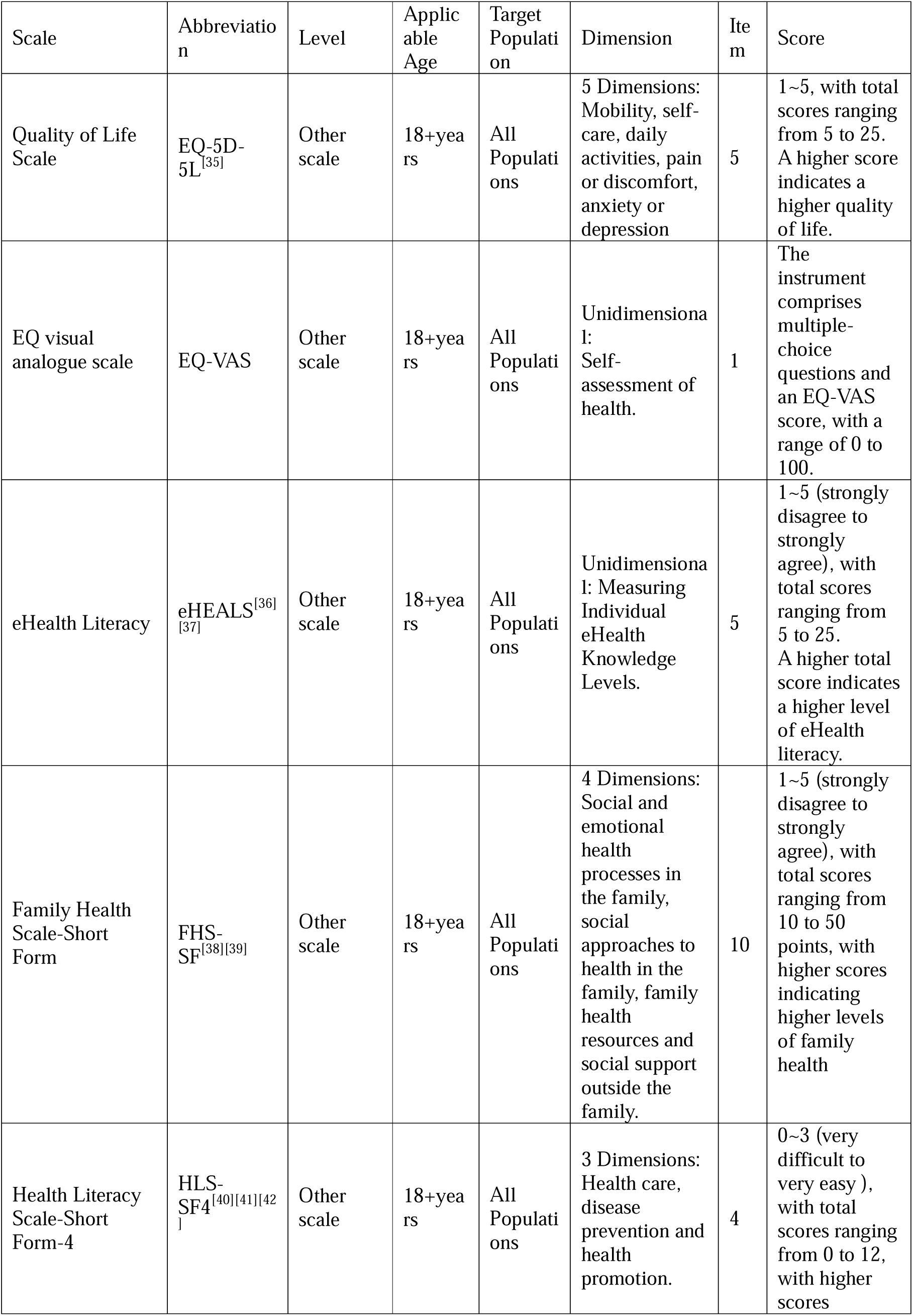
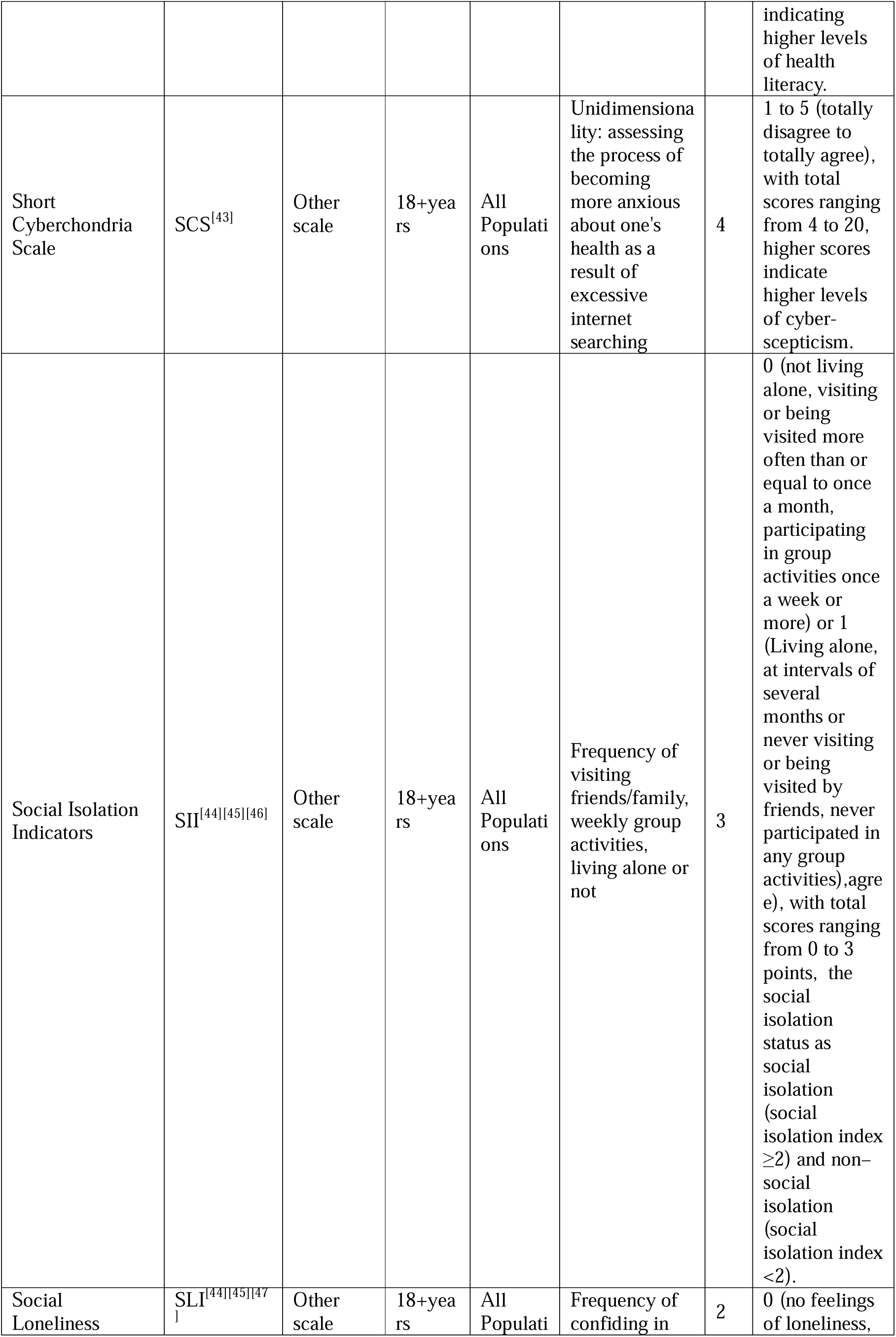
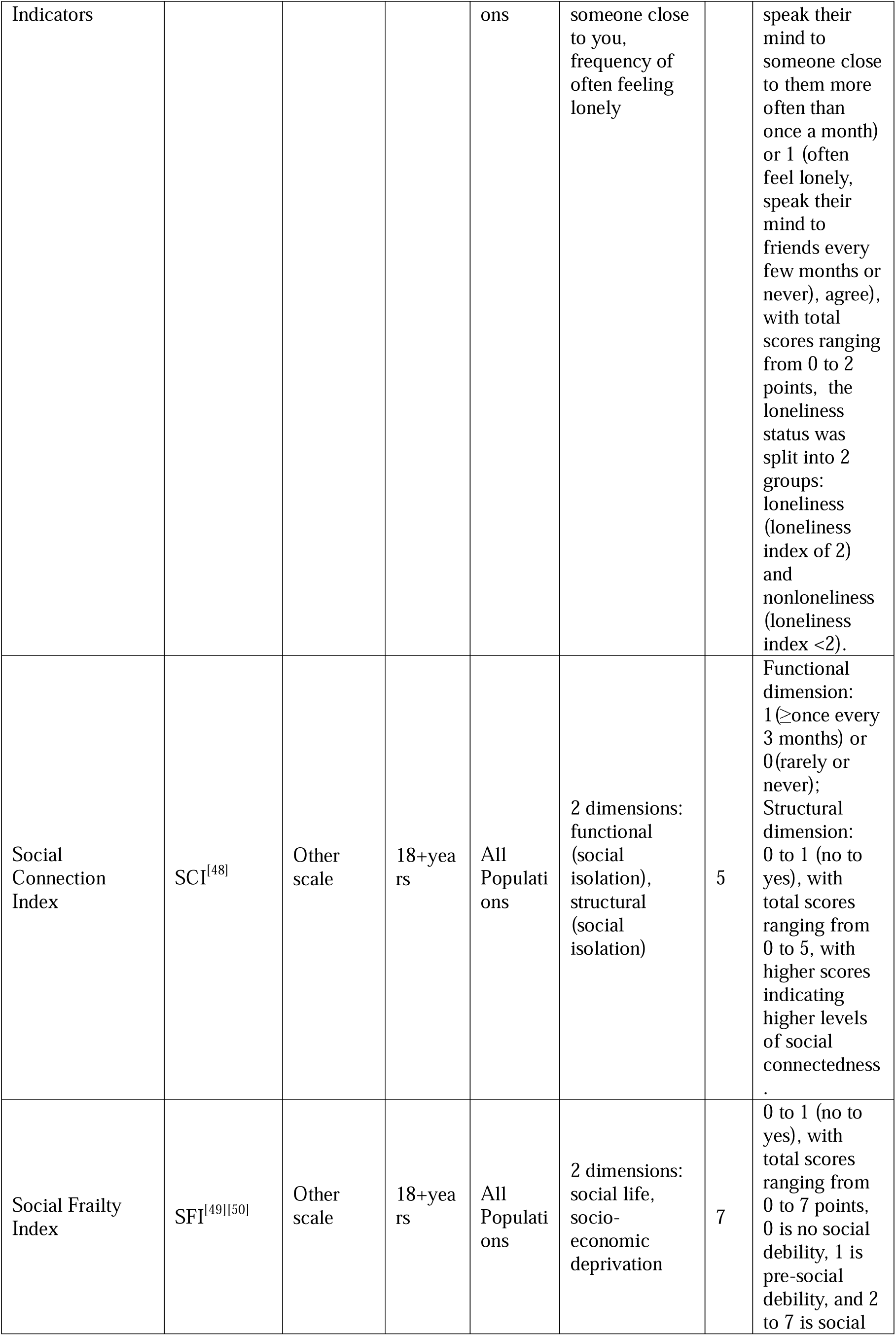
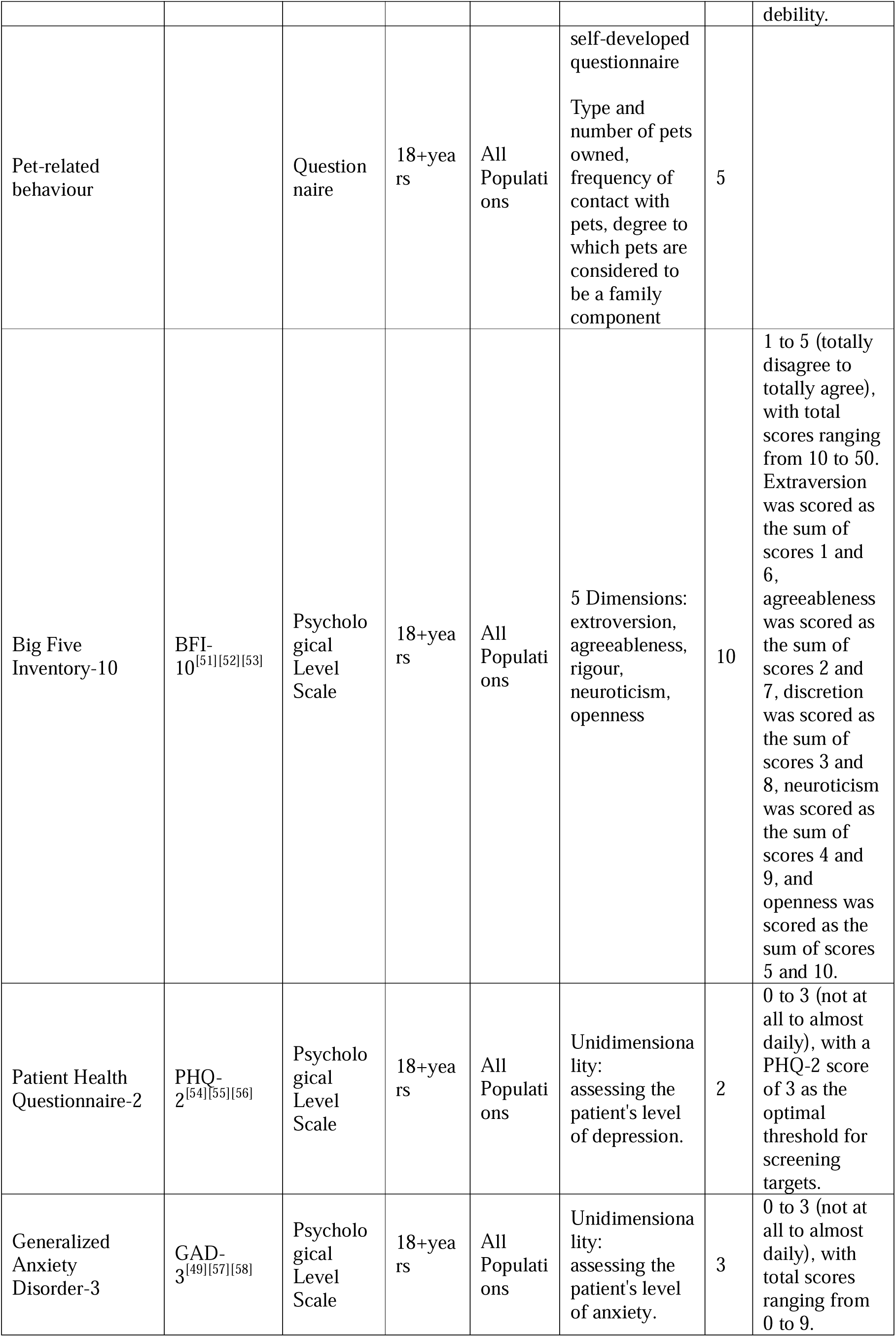
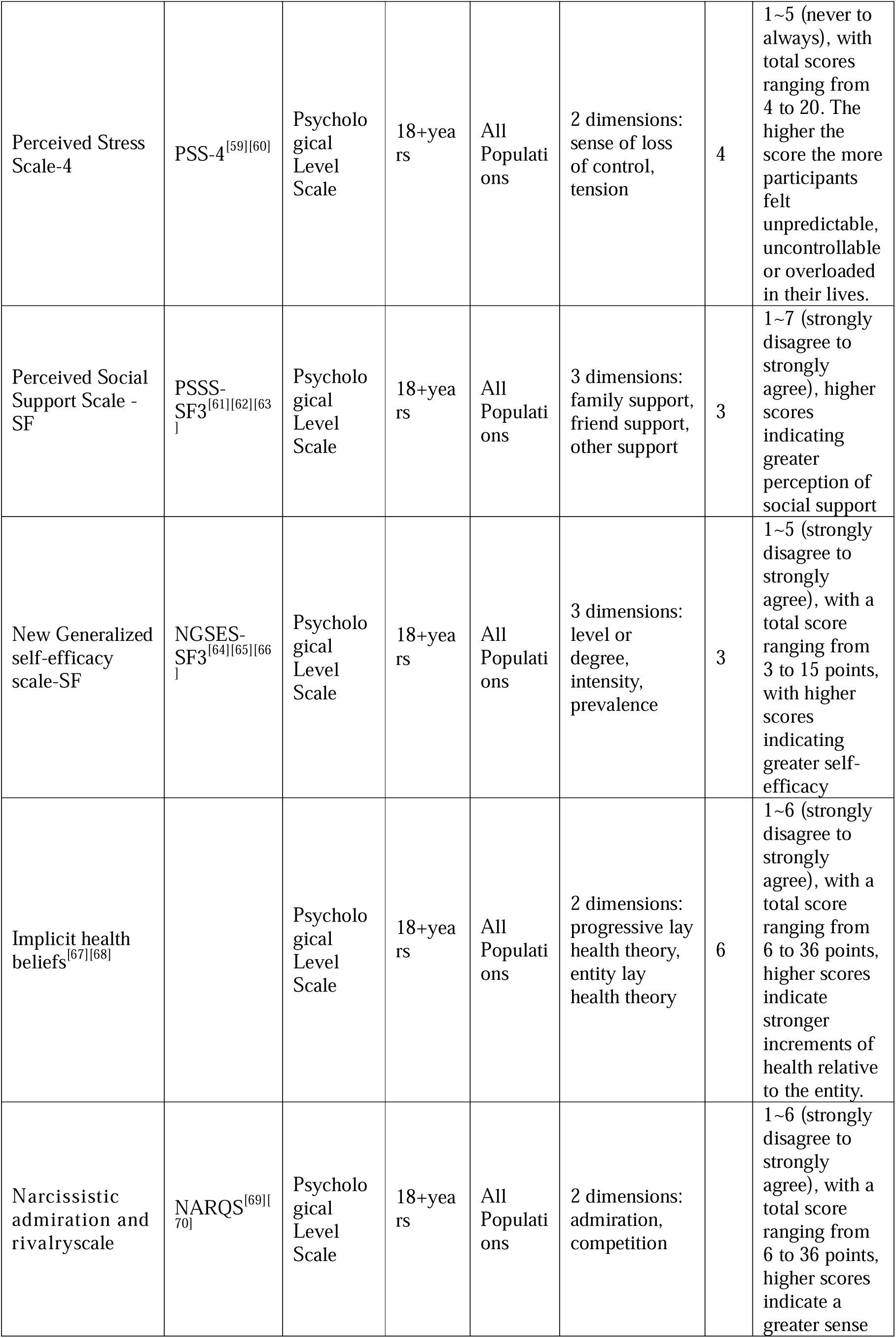
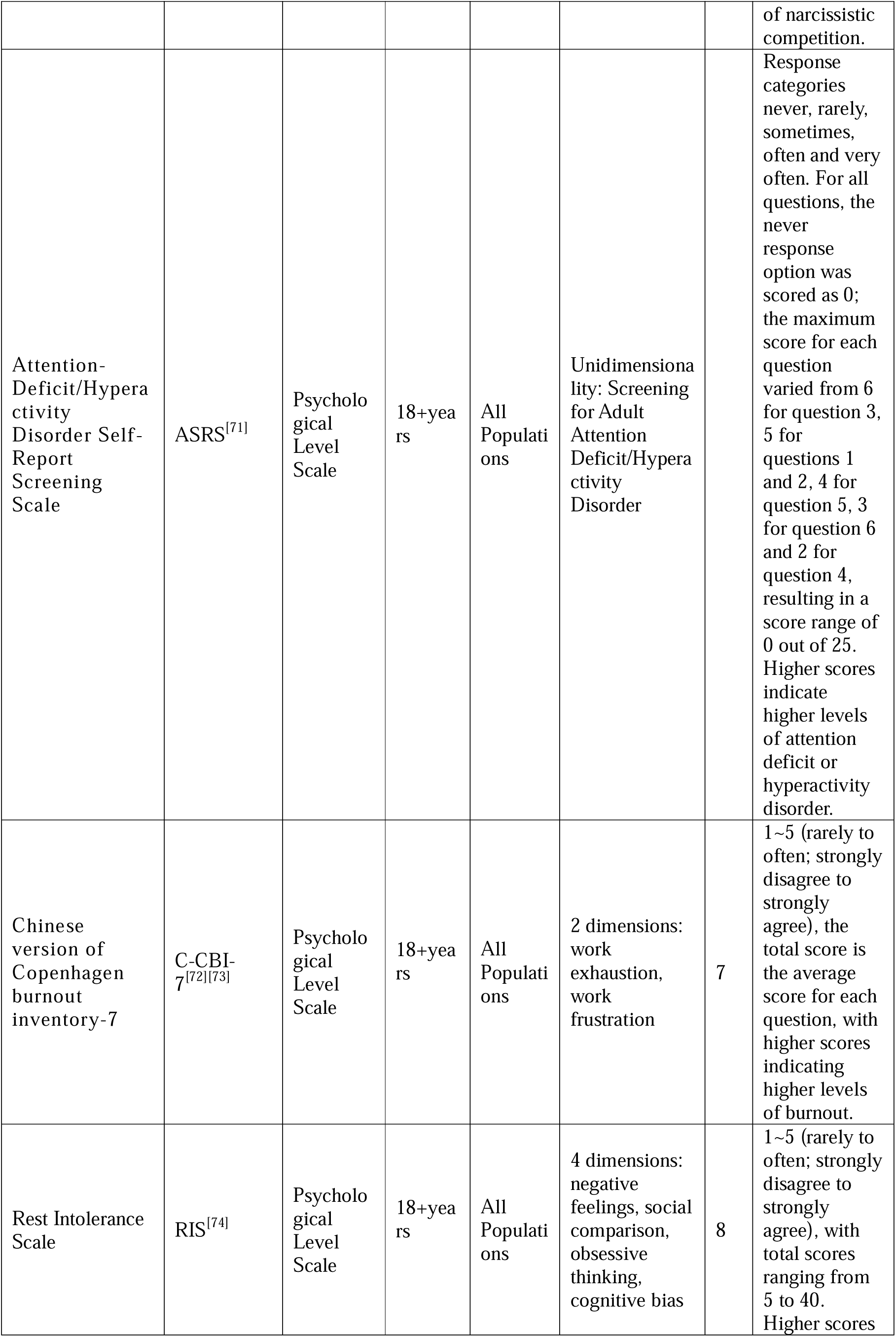
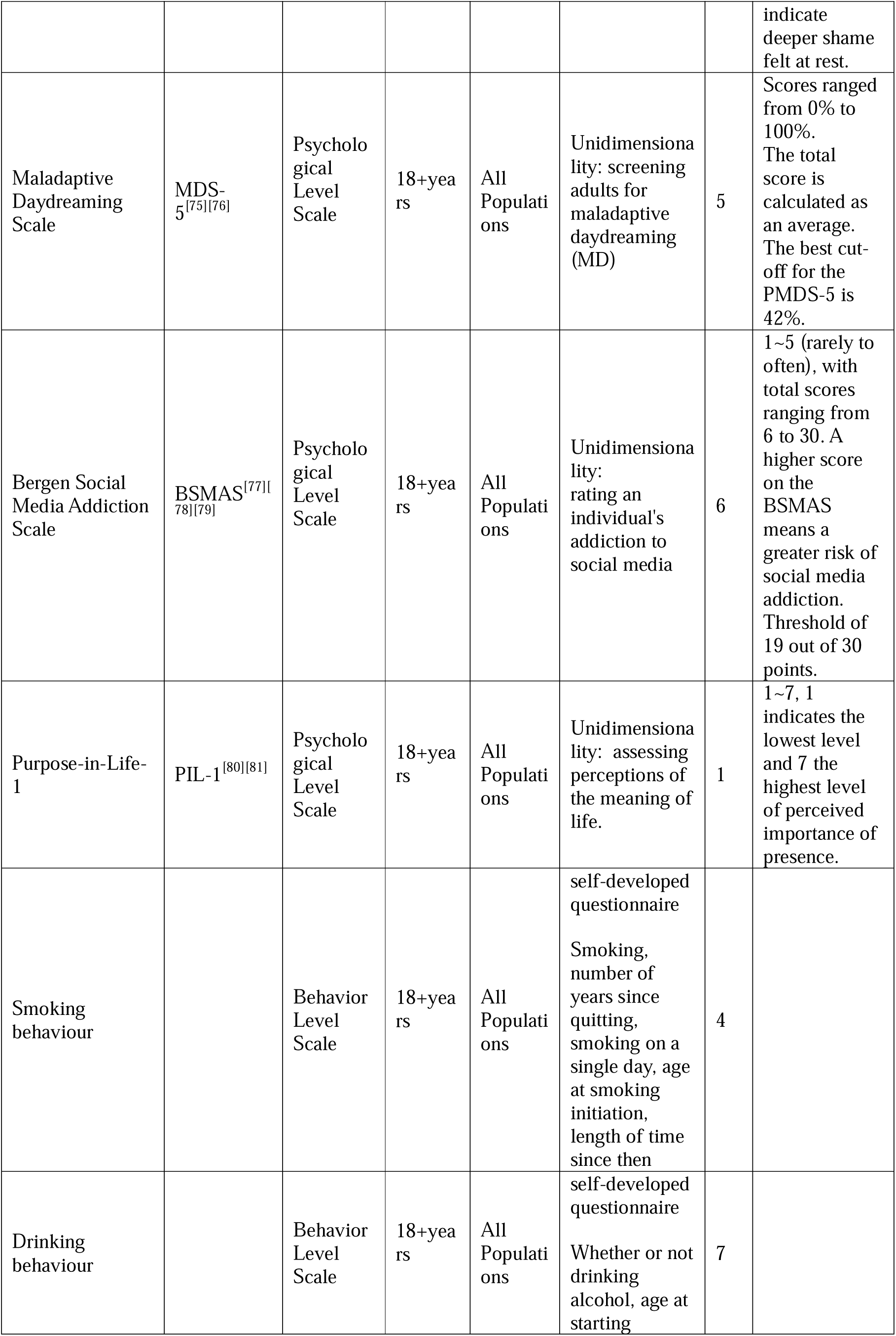
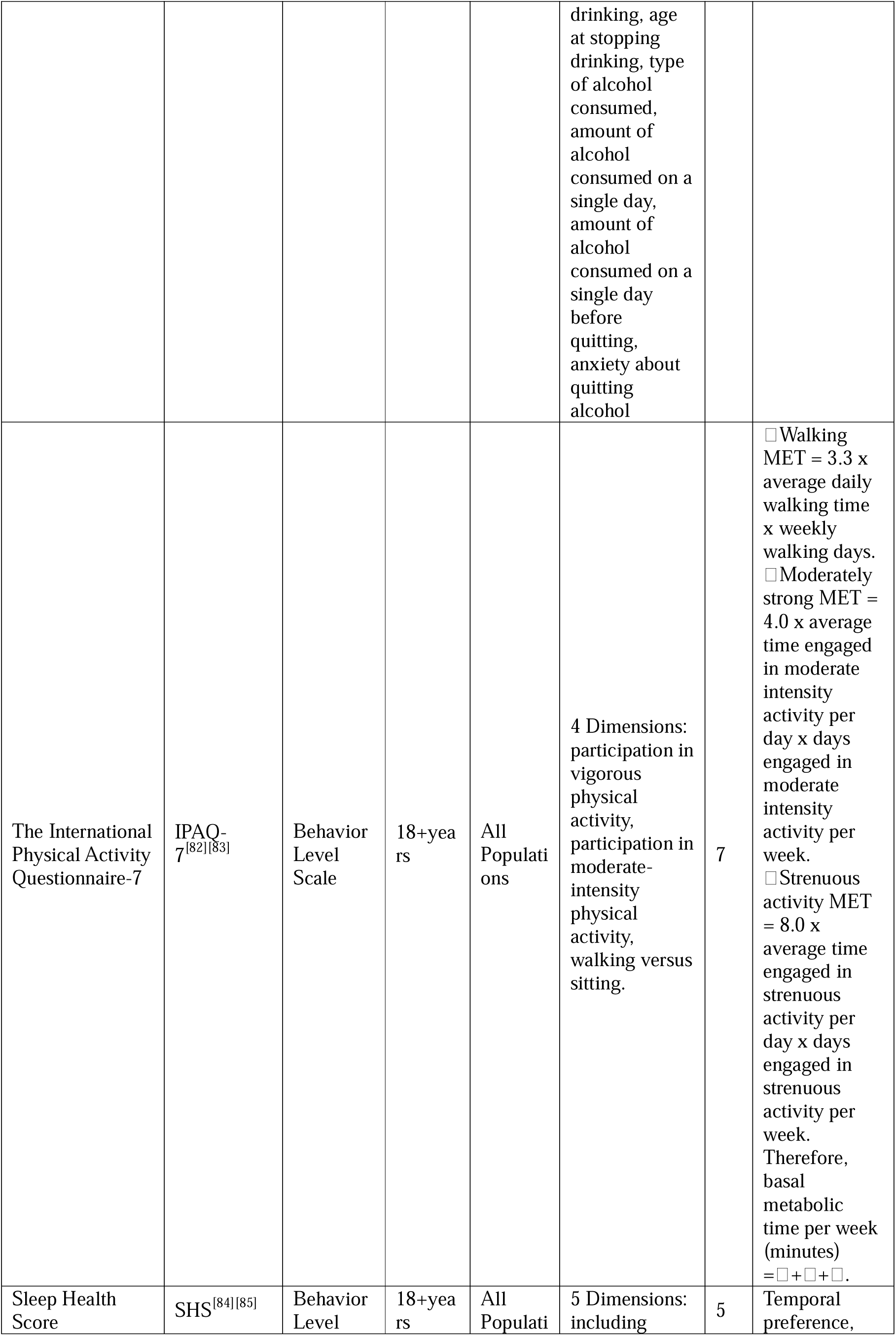
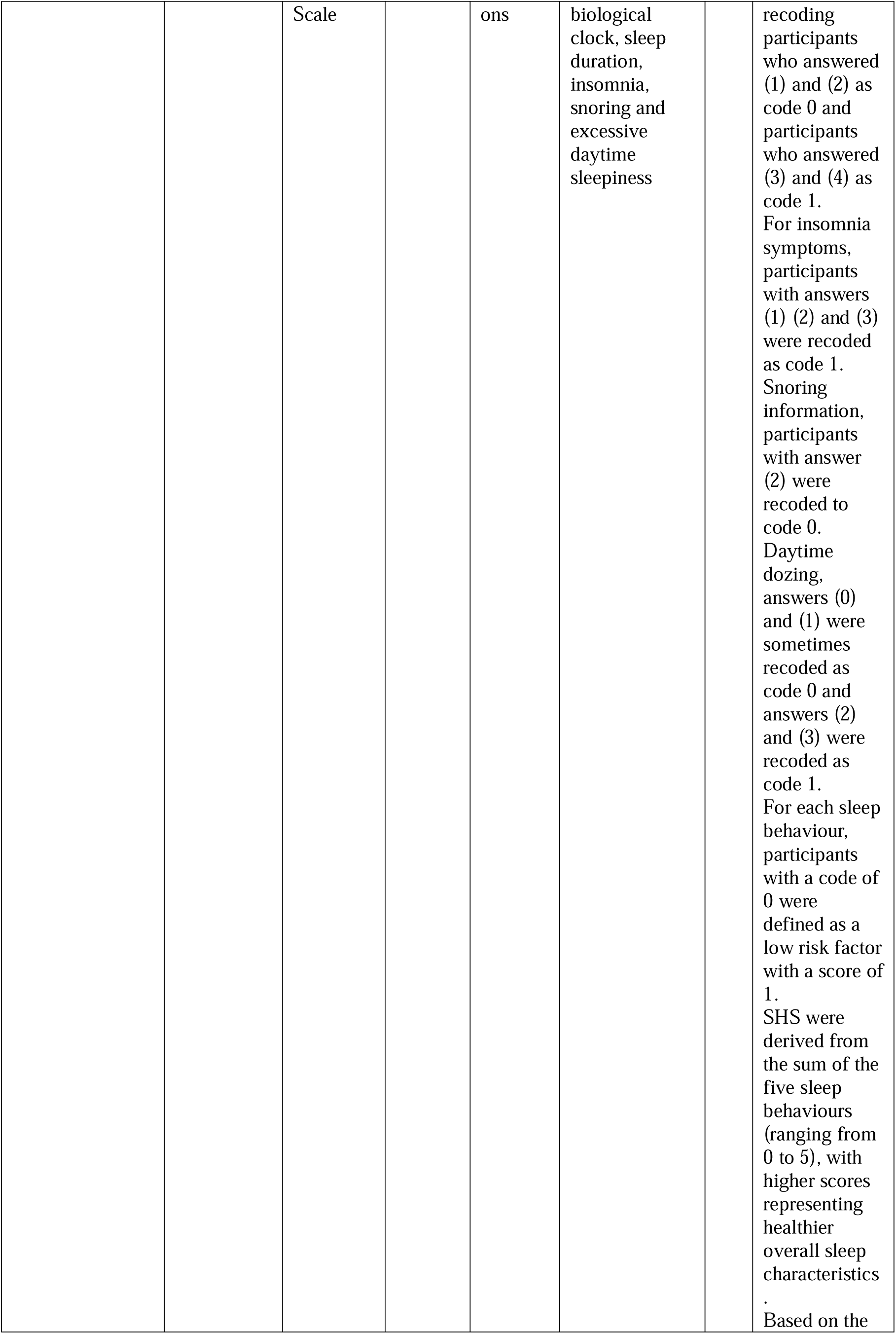
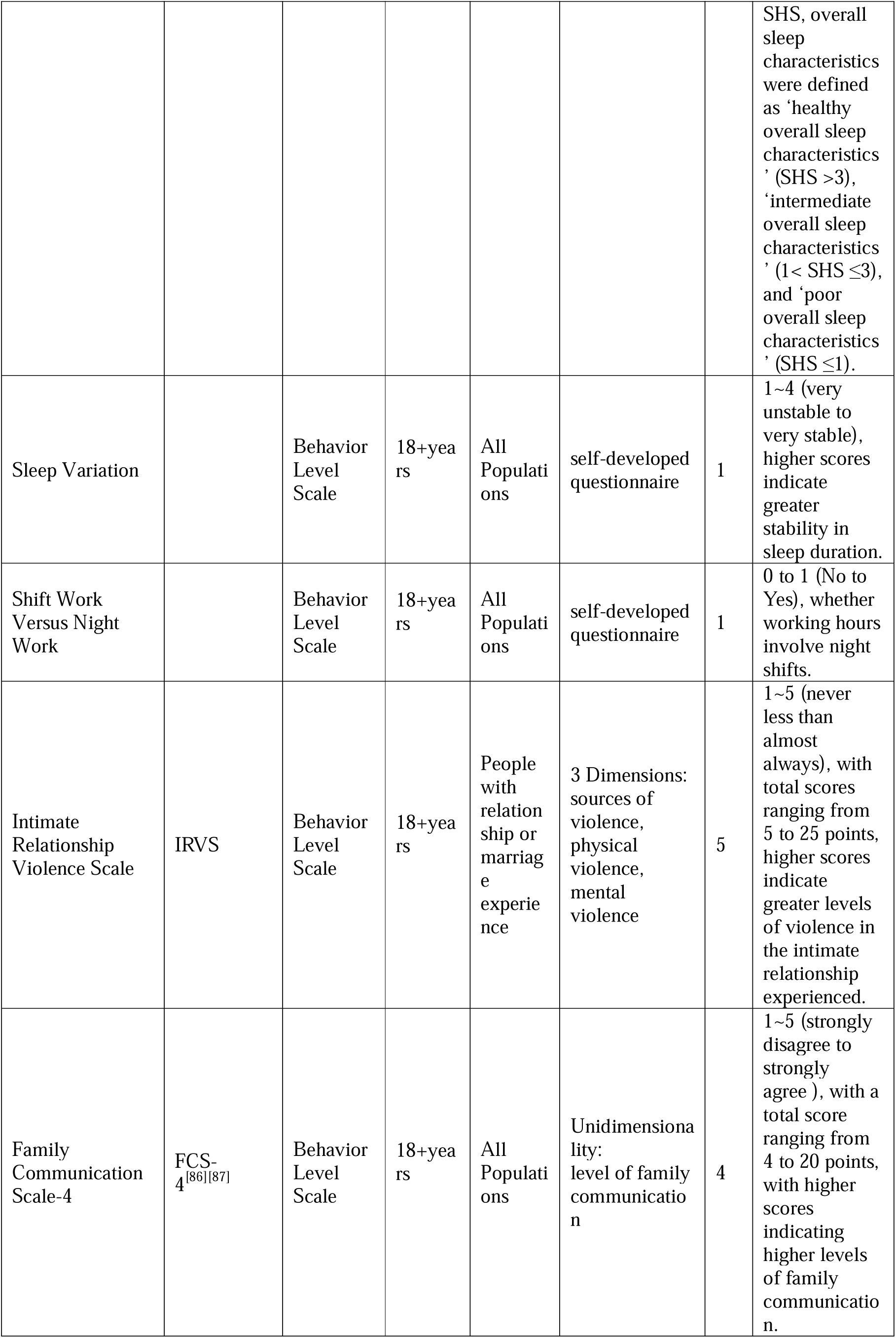
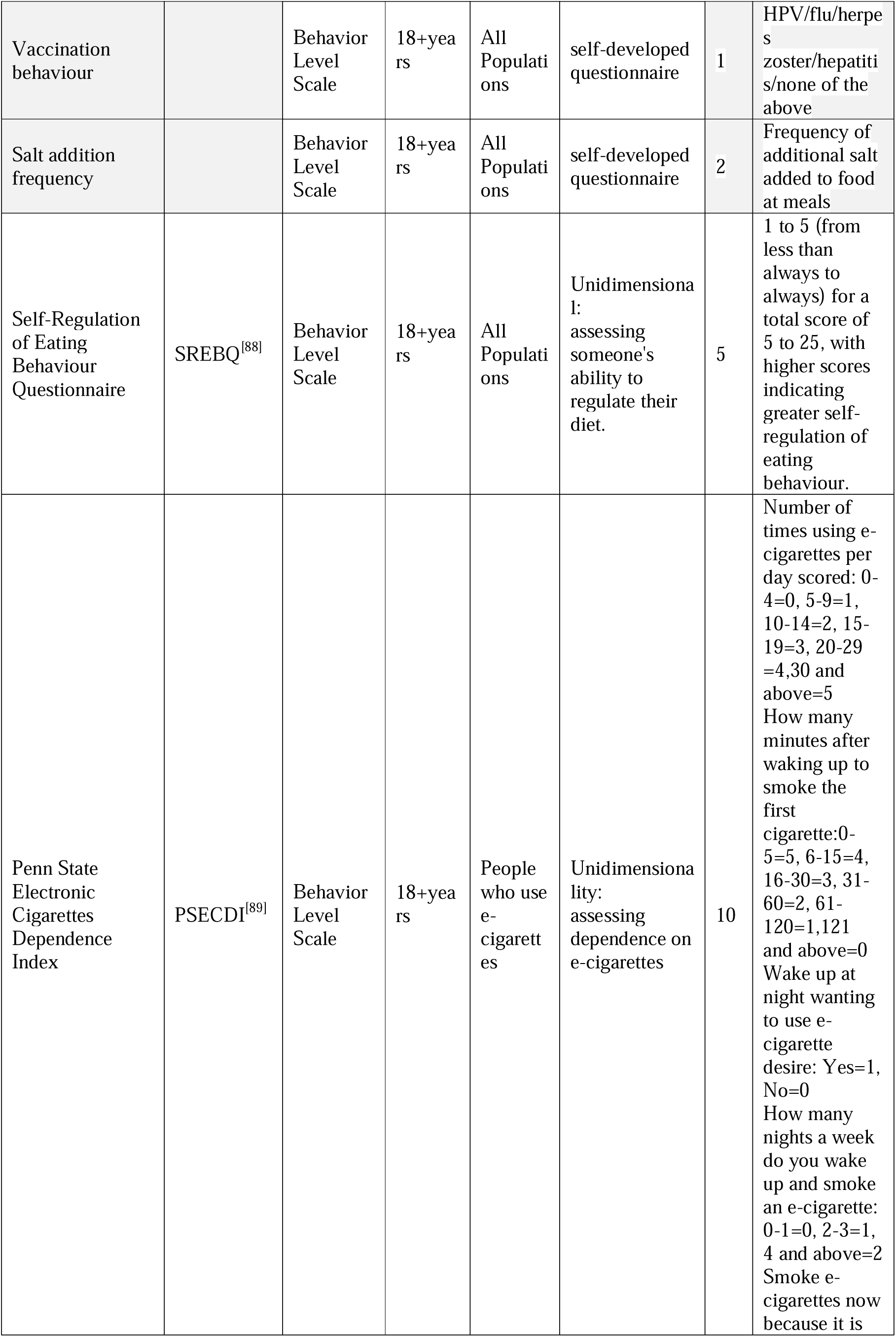
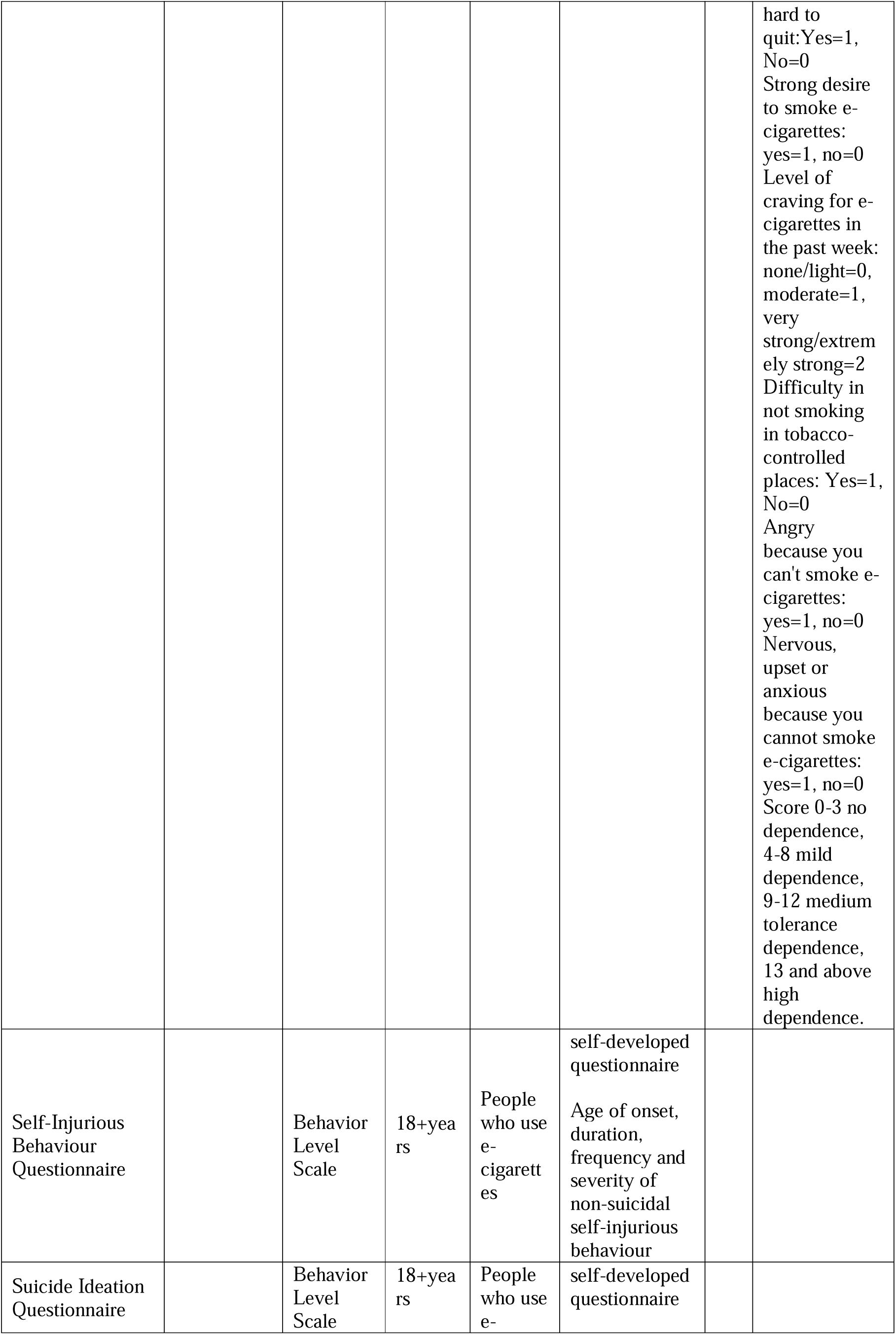
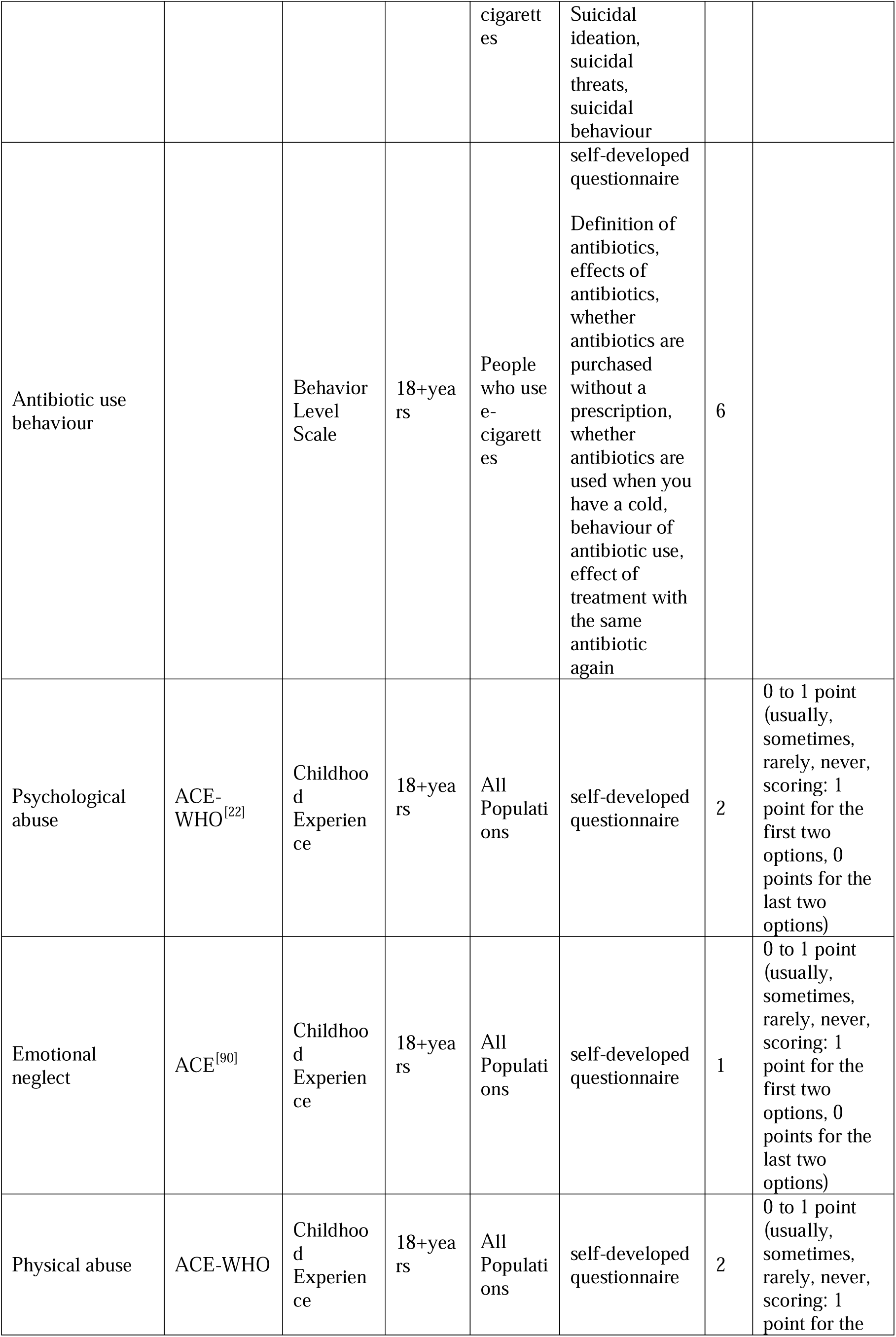
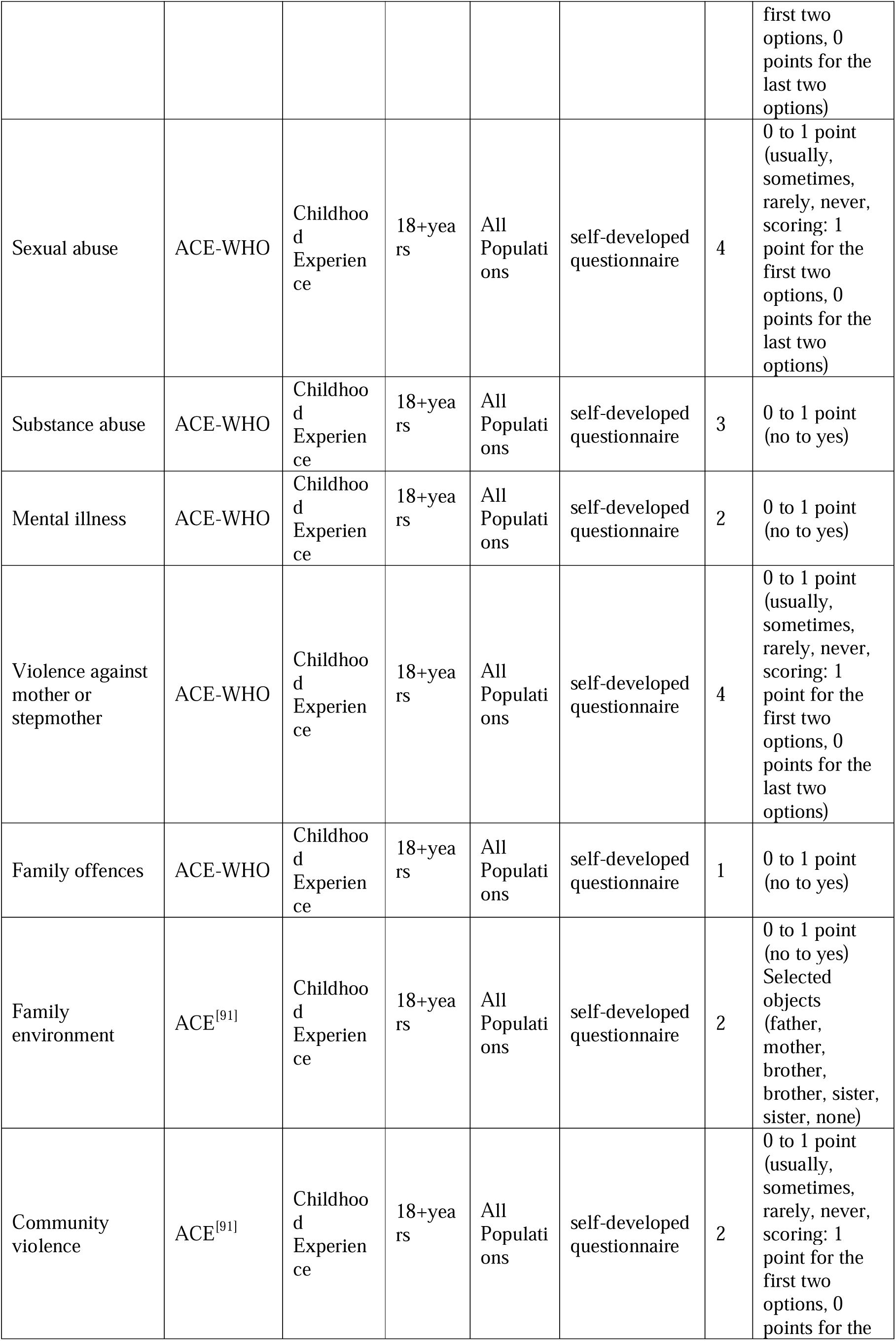
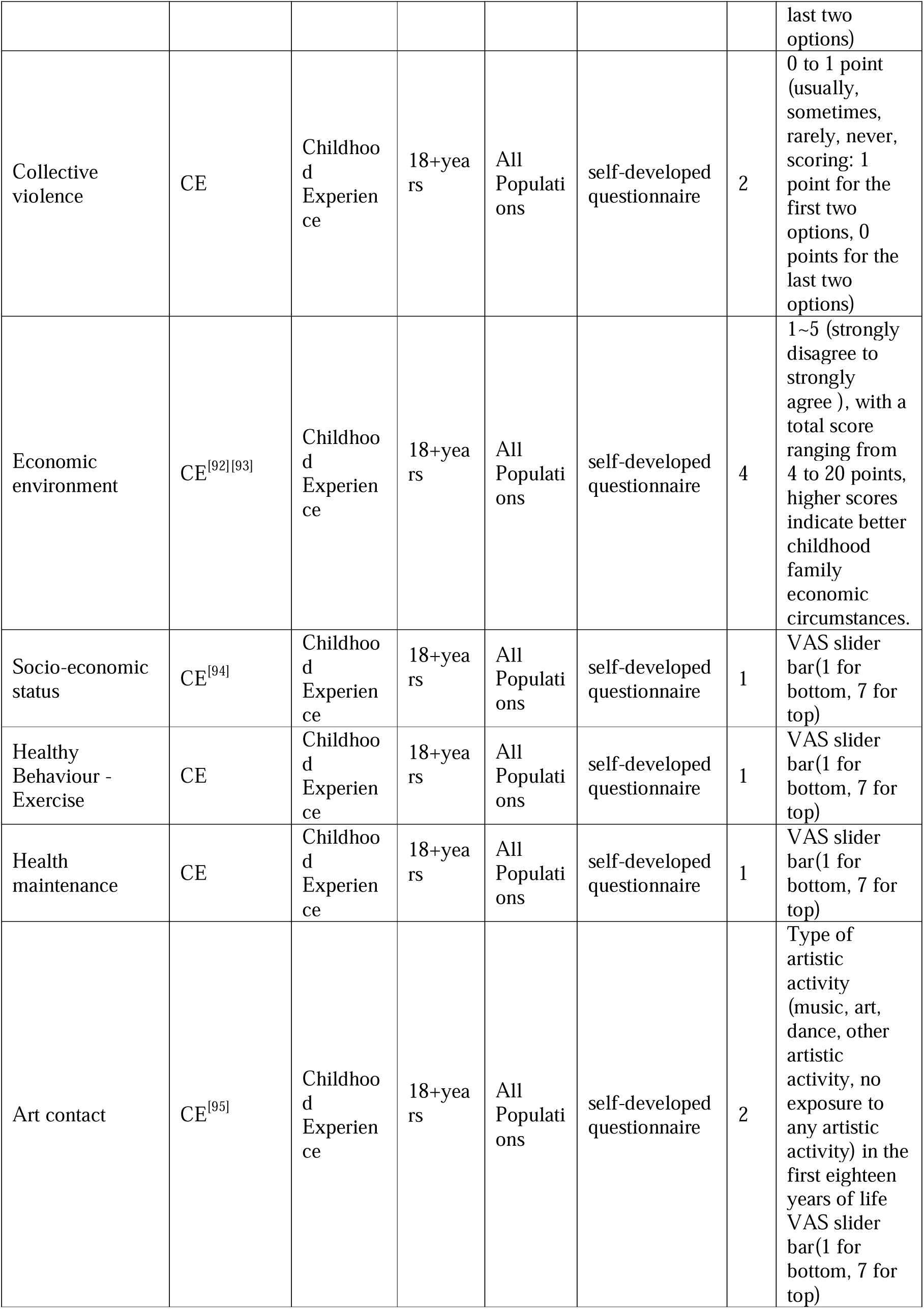
Scale used for the questionnaire.

#### 2.4.1 Basic personal information

This section comprises 21 questions on a range of topics, including biological sex at birth, sexual orientation, gender identity, ethnicity, religious beliefs, political status, employment status and current or pre-retirement employment, highest level of education attained, current period of study, academic background (school, faculty and major), relocation details (place of birth, registered residence, current place of residence, length of residence in current place of residence, last place and time of moving out), date of birth, and preferred hand.

#### 2.4.2 Personal Health Status

The Personal Health Status section comprises sixteen questions pertaining to the following: height, weight, waist circumference, chronic disease diagnosis, type of stroke, hypertension classification, hypertension-related complications, type of diabetes mellitus, classification of respiratory diseases, classification of urinary diseases, classification of gastrointestinal diseases, type of neoplasm, status of injuries, experience of life events, number of vaccinations and number of new crown infections.

#### 2.4.3 Household demographic information

This section comprises twenty-four questions pertaining to various aspects of the respondents’ household demographics. These include inquiries into the respondents’ family structure, their usual place of residence, the household registration, their marital status, their relationship history, their willingness to be classified as a ‘DINK’, their living arrangements (whether they live alone, with their spouse, with their spouse’s parents, or with their parents), and the number of their children. Furthermore, the respondents were asked whether they reside with their children, the age of the youngest and oldest children, the number of siblings, whether they reside with their siblings, the size of the dwelling, the dwelling structure, the number of properties owned, the debt status, the per capita monthly household income, the method of payment of medical expenses, financial issues related to medical payment, and the place of enrolment in basic health insurance.

#### 2.4.4 Socio-environmental situation

This section comprises three questions pertaining to the respondents’ neighbourhoods, social statuses, and whether they have enrolled in family doctor services.

#### 2.4.5 A hot topic in current affairs

The level of support for the integration of first aid knowledge into school health education, the level of support for the provision of free medical care for children under the age of five, the willingness to enrol one’s child in a childcare institution, the level of support for the implementation of a new national policy to permit unmarried women to freeze their eggs, the optimal timing for the delivery of life education, the perceived importance of life education for all, the acceptance of living wills, and the acceptance of palliative care, the level of support for the integration of first aid knowledge into school health education, The level of support for free medical care for children under five, the willingness to send one’s child to a childcare institution, the level of support for the introduction of a new national policy to allow unmarried women to freeze their eggs, the optimal time to provide life education, the importance of life education for all, the acceptance of living wills, and the acceptance of palliative care, hospice care, and the needs that exist during medical visits. The knowledge of the Sustainable Development Goals (SDGs); the utilisation of digital health interventions; the acceptance of digital health intervention modalities; the acceptance of immersive technologies (virtual reality, mixed reality, augmented reality) in healthcare; the acceptance of big language modelling in healthcare; the familiarity with carbon footprints; the familiarity with rare diseases; and the acceptance of the inclusion of high-value rare disease therapeutics in basic health insurance.

## 3 Discussion

The objective of this study is to provide a comprehensive and systematic picture of the current state of mental health and health behaviours of the Chinese population. To this end, a questionnaire has been designed with multidimensional variables and a large-scale cross-sectional survey has been implemented across multiple centres. It is anticipated that the findings of the study will provide robust data to inform a range of research areas and offer guidance to policy makers and healthcare organisations on how to implement changes to ensure the optimal well-being of the population and their families. The content of the survey will not only reveal the individual physical health status of Chinese residents and the impact of illness on their personal life and work, but also reflect their family status, medical conditions, childhood experiences, and economic and social network characteristics. In this study, questionnaires were employed to assess residents’ mental health status, perceived stress, and self-regulation ability. Additionally, the questionnaires were utilized to gain insight into residents’ substance use, daily activities, dietary habits, sleep quality, and social connections, as well as residents’ self-efficacy, rest shame, health beliefs, and family communication. The comprehensive scope and depth of the survey are sufficient to meet the need for in-depth knowledge of mental health and health behavior status.

In conducting the variable selection process, the researcher elected to retain approximately 30 per cent of the variables for comprehensive trajectory analysis. This was done to ensure that there were sufficient data points to enable the capture and understanding of key dynamics and trends in the data. The researchers aimed to analyse and interpret the complex patterns in the data in greater depth by retaining these variables, thereby facilitating the generation of more accurate and meaningful conclusions.

Furthermore, this study has a particular emphasis on individuals from specific backgrounds, with the objective of elucidating the influence of these background factors on their health and well-being. In order to achieve this objective, the research team constructed an extensive database containing data from a large number of questionnaires. Some of the data from the questionnaires were not only incorporated into the main database, but were also integrated into several sub-databases, including a cohort study for the maternal population. This design permits the comparison of data from different cohorts and facilitates cross-cohort analyses. In designing the questionnaire, the research team employed a range of validated scales for the assessment of participants’ health status. To illustrate, the Sleep Health Score is a scale that was initially derived from the UK BioBank study[12] and has been successfully applied to the China Kadoorie BioBank (CKB) cohort study[13]. The utilisation of these standardised scales will facilitate not only an in-depth data analysis within China, but also enable comparisons and alignments with international studies, thereby enhancing the global impact and application value of the study.

Based on rigorous field research and questionnaire design, this study constructed a survey database covering a wide range of psychological and behavioural characteristics of Chinese residents, and provided more comprehensive data support for a large sample. The findings are intended to provide guidance to policy makers and health care organisations to promote the improvement of the physical and mental health of Chinese citizens. Between 2021 and 2023, the research team conducted a cross-sectional survey on the mental and behavioural health of Chinese families, which yielded significant findings[14][15]. Looking forward to 2024, the study expects to further contribute to the health of the Chinese people and people around the world. This year’s study has established a committee of investigators based on previous years, on which a more stringent quality control process and management framework has been developed. During the distribution and collection of questionnaires, members of the Working Committee, comprising the Secretary General, the Deputy Secretary General and the Secretary, were responsible for the management functions to ensure strict control over the underlying logic and quality control of the database. The publication of this research programme enhances the transparency of the research and informs the scientific community of ongoing research, which helps to avoid duplication of research efforts and promotes coordination of research efforts.

The ‘Social Phenomena’ section of the study presents a comprehensive and detailed analysis of a number of variables, including identify flow population, rest intolerance and sense of meaning in life. By undertaking a detailed examination of these variables, the researchers were able to gain a more comprehensive understanding of the complex factors that underpin this social phenomenon. This, in turn, provides a more scientific and precise basis for decision-making and policy planning in related fields. In particular, the researcher’s identification of mobile populations enables the assessment of inter-regional economic, social, and cultural ties, the optimisation of resource allocation, and the improvement of the effectiveness of public services[16]. Secondly, the researchers conducted a detailed study on rest tolerance[17], analysing the coping strategies and psychological mechanisms of different populations in the face of stress and fatigue. This provided a basis for the formulation of more effective rest and recovery policies. Finally, the researcher evaluates the perception of meaning in life, which helps to analyse how people search for and construct meaning in life in modern society, and the impact of this process on social stability and individual psychological health[18]. By comprehensively analysing these key variables, the researcher not only reveals the deep-seated reasons behind social phenomena, but also provides policy makers with a more scientific and precise basis for decision-making, thus promoting the harmonious development of society.

Furthermore, this study conducted a comprehensive examination of the prevalence of occupational health concerns, e-cigarette addiction, and antibiotic use behaviours, which have emerged as significant issues in the context of the contemporary fast-paced lifestyle. A deeper comprehension of these variables will facilitate the elucidation of the potential influence of occupational health concerns, e-cigarette addictive behaviours and antibiotic use habits on the health status of individuals in the context of modern, fast-paced lifestyles. Furthermore, it will enable the investigation of the mechanisms through which these effects are manifested. Firstly, in the context of the fast-paced modern life, it can be posited that work-related stress and occupational issues may have a significant impact on individuals’ mental health and behavioural patterns. By evaluating occupational health, researchers can ascertain the influence of the work environment and occupational stress on an individual’s health behaviours[19], including dietary habits, sleep quality, and exercise frequency. This allows for the elucidation of how occupational factors can act as drivers of behavioural change in a fast-paced lifestyle[20]. Secondly, e-cigarette dependence may be related to the coping strategies and psychological mechanisms of individuals in a fast-paced life[21], as it constitutes a behavioural pattern. The assessment of e-cigarette dependence levels may provide a scientific basis for policy makers to consider the impact of a fast-paced lifestyle on individuals’ behaviours in public health policies and practices. This could facilitate the formulation of more effective preventive and intervention measures. Finally, antibiotic use is closely related to an individual’s health status and medical behaviours, which can reveal potential health risks for individuals in a fast-paced lifestyle[22]. The assessment of antibiotic use can assist policy makers and health care organisations in understanding and responding to the challenges posed by fast-paced lifestyles. By examining these aspects in detail, this study can provide robust data and a scientific basis for the development and optimisation of relevant health interventions. Furthermore, it can guide policymakers to consider the impact of fast-paced lifestyles on individual behaviours in public health policies and practices, thus facilitating improvements in people’s health, a reduction in the incidence of adverse health behaviours, and an enhancement of the overall level of public health.

The study special focus on harm-related variables, including childhood experiences (adverse and favourable experiences, as well as upbringing), self-injurious behaviours, and a comprehensive assessment of the level of many variables related to socialisation, such as social isolation, social bonding, and social loneliness. Additionally, the study places a particular emphasis on the functioning of pets.

In examining the analysis of variables related to harm, it is essential to consider the individual’s experiences during childhood[23], encompassing both adverse and favourable experiences, as well as the various factors that shape the process of growing up. The period of childhood is a crucial phase in the development of an individual’s personality and behavioural patterns. Consequently, the early environments and experiences a person encounters have a significant influence on their mental health and behavioural tendencies. Adverse childhood experiences may include exposure to domestic violence, neglect, abuse, or other forms of trauma. These experiences can give rise to a range of psychological and behavioural issues in adulthood[24]. Conversely, positive childhood experiences, such as the provision of familial support, care and a positive social environment, can provide individuals with a robust psychological foundation that enables them to better cope with future challenges.

In addition to childhood experiences, self-harming behaviours represent a significant area of concern[25]. Self-injurious behaviours may manifest in a variety of forms, including cutting, burning, or other self-harming actions. These behaviours are frequently a means of coping with internal distress and stress. The underlying causes of self-harming behaviours frequently encompass profound psychological issues, including depression, anxiety, and personality disorders. Consequently, it is imperative to comprehend and examine the underlying motives behind these behaviours in order to prevent and address them effectively.

It is also imperative to consider the role of social isolation, social connectedness and social loneliness in psychological and behavioural mechanisms. The term “social isolation” is used to describe individuals who perceive themselves to be isolated from their social relationships and who lack the social support and connections that they require[26]. Prolonged social isolation has been linked to an increased risk of psychological and emotional problems, as well as an elevated likelihood of suicidal ideation and behaviour. Conversely, positive social connections and networks can provide emotional support, enhance psychological resilience, and facilitate more effective coping with life’s stresses and challenges. In contrast, social isolation can be defined as an individual’s perception of being isolated and unappreciated within their social relationships. This can give rise to psychological distress and negative emotional states[27].

It is notable that this study posits that the functional role of pets should not be overlooked[28]. In addition to providing emotional support and companionship, pets have been demonstrated to alleviate feelings of loneliness and stress[29]. It has been demonstrated that interaction with animals can result in a reduction in blood pressure, an alleviation of anxiety and depressive symptoms, and an improvement in socialisation. It can therefore be argued that pets play an important role in an individual’s mental health and emotional regulation.

In conclusion, a number of factors, including childhood experiences, self-injurious behaviours, social isolation/social bonding/social loneliness, and the functional role of pets, have been considered. By undertaking a comprehensive examination of these elements, we can gain a deeper insight into the psychological state and behavioural patterns of individuals, thereby establishing a scientific foundation for the development of prevention and intervention strategies.

Furthermore, in light of the dynamic nature of China’s policies, this study has incorporated an investigation of the aspirations pertaining to egg freezing for single women[30] (with the objective of fostering childbirth and curbing population growth), palliative care[31], death education[32], and advance care directives into the Hot Topics in Society section. The objective of these additions is to provide a more comprehensive reflection of the current social issues and to examine the social, ethical and legal implications of these issues in greater depth. It is our intention that the study of these issues will provide useful references and suggestions for the formulation and implementation of relevant policies.

In order to proactively address the challenges of the Global Health Goals, this study has been expanded this year to include several new and important topics related to climate change and mental disorders. The topics under examination include the impact of extreme weather events[33] on mental health and the extent to which carbon footprints[34] are understood, with a view to exploring the ways in which these environmental issues interact with human mental health. Furthermore, the research team investigated the acceptance of data science applications in healthcare, with a particular focus on the potential of artificial intelligence technologies in disease diagnosis, treatment and prevention. Furthermore, the study incorporated virtual reality technology, an emerging tool, in addition to assessing the role that digital health interventions, such as mobile health applications and online psychosocial support platforms, can play in promoting public health. It is our intention that the results of these extended studies will provide a more comprehensive and in-depth scientific basis and practical guidance for achieving global health goals.

## Data Availability

All data produced in the present study are available upon reasonable request to the authors

## Author contributions

Writing – Original Draft Preparation, S.F&Y.L&F.D&W.C&Q.H&T.D; Writing – Review & Editing, J.Z&Q.S&Y.B&Y.L&S.Z&X.Z&F.J&W.M&X.S&Y.W; Data Curation, S.F&F.D; Supervision, X.Z&F.J&W.M&Y.W; Study Deisgn & Supervision & Funding Acquisition & Resources, X.S&Y.W; Project Administration, S.F&W.C&Q.H.

## Funding

This project was established with the Key Laboratory of Health Economics and Policy Research of the National Health Commission(NHC-HEPR202401).

## Availability of data and materials

The PBICR project has been ongoing from 2020 to the present, with an annual multi center extensive sample size transect survey. We usually distinguish between different years of surveys by the suffix (e.g., PBICR2024 denotes surveys conducted by PBICR in 2024). The historical materials and any associated protocols supporting this study’s findings are available from the corresponding authors upon request.

## Declarations

### Research ethics and patient consent

This study (H20240237I) has been approved by the SHANGHAI JIAOTONG UNIVERSITY Institutional Review Board for Human Research Protections. The cover page of the questionnaire will explicitly outline the study’s objectives and ensure the preservation of anonymity, confidentiality, and the participants’ right to decline participation. Informed consent has been duly acquired from all individuals involved in the study. This study was filed in the National Health Security Information Platform (Record No.: MR-37-23-017876) and officially registered in the China Clinical Trials Registry(Registration No.: ChiCTR2400085016).

## Acknowledgments

Not applicable.

## Consent for publication

Not applicable. Availability of data and material.

## Competing interests

All authors declare that they have no competing interests.

## Abbreviation

COVID-19: Corona Virus Disease 2019
PBICR: Psychology and Behavior Investigation of Chinese Residents
WHO: World Health Organization

## Notes

### Competing Interest Statement

The authors have declared no competing interest.

### Author Declarations

This study (H20240237I) has been approved by the SHANGHAI JIAOTONG UNIVERSITY Institutional Review Board for Human Research Protections. The cover page of the questionnaire will explicitly outline the study's objectives and ensure the preservation of anonymity, confidentiality, and the participants' right to decline participation. Informed consent has been duly acquired from all individuals involved in the study.

## References

[1] McGrath JJ, Al-Hamzawi A, Alonso J, Altwaijri Y, Andrade LH, Bromet EJ, Bruffaerts R, de Almeida JMC, Chardoul S, Chiu WT, Degenhardt L, Demler OV, Ferry F, Gureje O, Haro JM, Karam EG, Karam G, Khaled SM, Kovess-Masfety V, Magno M, Medina-Mora ME, Moskalewicz J, Navarro-Mateu F, Nishi D, Plana-Ripoll O, Posada-Villa J, Rapsey C, Sampson NA, Stagnaro JC, Stein DJ, Ten Have M, Torres Y, Vladescu C, Woodruff PW, Zarkov Z, Kessler RC; WHO World Mental Health Survey Collaborators. Age of onset and cumulative risk of mental disorders: a cross-national analysis of population surveys from 29 countries. Lancet Psychiatry. 2023 Sep;10(9):668–681. doi: 10.1016/S2215-0366(23)00193-1. Epub 2023 Jul 30. PMID: 37531964; PMCID: PMC10529120.

[2] Li W, Yang Y, Liu ZH, Zhao YJ, Zhang Q, Zhang L, Cheung T, Xiang YT. Progression of Mental Health Services during the COVID-19 Outbreak in China. Int J Biol Sci. 2020 Mar 15;16(10):1732–1738. doi: 10.7150/ijbs.45120. PMID: 32226291; PMCID: PMC7098037.

[3] Freeman M. The World Mental Health Report: transforming mental health for all. World Psychiatry. 2022 Oct;21(3):391–392. doi: 10.1002/wps.21018. PMID: 36073688; PMCID: PMC9453907.

4. World Health Organization. (2024). Behavioural Sciences for Better Health. https://www.who.int/initiatives/behavioural-sciences

[5] Viner R, Russell S, Saulle R, Croker H, Stansfield C, Packer J, Nicholls D, Goddings AL, Bonell C, Hudson L, Hope S, Ward J, Schwalbe N, Morgan A, Minozzi S. School Closures During Social Lockdown and Mental Health, Health Behaviors, and Well-being Among Children and Adolescents During the First COVID-19 Wave: A Systematic Review. JAMA Pediatr. 2022 Apr 1;176(4):400–409. doi: 10.1001/jamapediatrics.2021.5840. PMID: 35040870.

[6] Wu Y, Fan S, Liu D, Sun X. Psychological and behavior investigation of Chinese residents: Concepts, practices, and prospects. Chinese General Practice Journal. 2024; 1(3): 149–156. doi: 10.1016/j.cgpj.2024.07.006.

[7] Lim DC, Najafi A, Afifi L, Bassetti CLA, Buysse DJ, Han F, Högl B, Melaku YA, Morin CM, Poyares D, Somers VK, Eastwood PR, Zee PC, Jackson CL. The need to promote sleep health in public health agendas across the globe. Lancet Public Health. 2023 Oct;8(10):e820–e826. doi: 10.1016/S2468-2667(23)00182-2.

[8] McCloud T, Kamenov S, Callender C, Lewis G, Lewis G. The association between higher education attendance and common mental health problems among young people in England: evidence from two population-based cohorts. Lancet Public Health. 2023 Oct;8(10):e811–e819. doi: 10.1016/S2468-2667(23)00188-3. PMID: 37777290; PMCID: PMC10958987.

[9] Campbell F, Blank L, Cantrell A, Baxter S, Blackmore C, Dixon J, Goyder E. Factors that influence mental health of university and college students in the UK: a systematic review. BMC Public Health. 2022 Sep 20;22(1):1778. doi: 10.1186/s12889-022-13943-x. PMID: 36123714; PMCID: PMC9484851.

[10] Wilhite K, Booker B, Huang BH, Antczak D, Corbett L, Parker P, Noetel M, Rissel C, Lonsdale C, Del Pozo Cruz B, Sanders T. Combinations of Physical Activity, Sedentary Behavior, and Sleep Duration and Their Associations With Physical, Psychological, and Educational Outcomes in Children and Adolescents: A Systematic Review. Am J Epidemiol. 2023 Apr 6;192(4):665–679. doi: 10.1093/aje/kwac212. PMID: 36516992; PMCID: PMC10089066.

[11] Yang Y, Fan S, Chen W, Wu Y. Broader Open Data Needed in Psychiatry: Practice from the Psychology and Behavior Investigation of Chinese Residents. Alpha Psychiatry. 2024 Aug 1;25(4):564–565. doi: 10.5152/alphapsychiatry.2024.241804. PMID: 39360297; PMCID: PMC11443289.

[12] Yao Y, Jia Y, Wen Y, Cheng B, Cheng S, Liu L, Yang X, Meng P, Chen Y, Li C, Zhang J, Zhang Z, Pan C, Zhang H, Wu C, Wang X, Ning Y, Wang S, Zhang F. Genome-Wide Association Study and Genetic Correlation Scan Provide Insights into Its Genetic Architecture of Sleep Health Score in the UK Biobank Cohort. Nat Sci Sleep. 2022 Jan 6;14:1–12. doi: 10.2147/NSS.S326818. PMID: 35023977; PMCID: PMC8747788.

[13] Fan M, Sun D, Zhou T, Heianza Y, Lv J, Li L, Qi L. Sleep patterns, genetic susceptibility, and incident cardiovascular disease: a prospective study of 385-292 UK biobank participants. Eur Heart J. 2020 Mar 14;41(11):1182–1189. doi: 10.1093/eurheartj/ehz849. PMID: 31848595; PMCID: PMC7071844.

[14] Zhang X, Zhang X, Li Y, Chen T, Wang Y, Siow L, Wang Y, Ming W-K, Sun X, Wu Y. Factors affecting acceptance of palliative care in mainland China: a national cross-sectional study. Lancet Oncol. 2022. doi: 10.1016/S1470-2045(22)004028.

[15] Bao Y, Wang C, Xu H, Lai Y, Yan Y, Ma Y, Yu T, Wu Y. Effects of an mHealth Intervention for Pulmonary Tuberculosis Self-management Based on the Integrated Theory of Health Behavior Change: Randomized Controlled Trial. JMIR Public Health Surveill. 2022 Jul 14;8(7):e34277. doi: 10.2196/34277. PMID: 35834302; PMCID: PMC9335179.

[16] Horn AL, Bell BM, Bulle Bueno BG, Bahrami M, Bozkaya B, Cui Y, Wilson JP, Pentland A, Moro E, de la Haye K. Population mobility data provides meaningful indicators of fast food intake and diet-related diseases in diverse populations. NPJ Digit Med. 2023 Nov 15;6(1):208. doi: 10.1038/s41746-023-00949-x. PMID: 37968446; PMCID: PMC10651929.

[17] Wang F, Song H, Meng X, Wang T, Zhang Q, Yu Z, Fan S, Wu Y. Development and validation of the long and short forms of the rest intolerance scale for college students. Personality and Individual Differences. 2025;233:112869. ISSN 0191-8869. doi: 10.1016/j.paid.2024.112869.

[18] Chen R, Liu YF, Huang GD, Wu PC. The relationship between physical exercise and subjective well-being in Chinese older people: The mediating role of the sense of meaning in life and self-esteem. Front Psychol. 2022 Nov 10;13:1029587. doi: 10.3389/fpsyg.2022.1029587. PMID: 36438332; PMCID: PMC9685655.

[19] González-Caballero J. Occupational health nursing: Realities and challenges. Int Nurs Rev. 2024 Sep;71(3):513–520. doi: 10.1111/inr.12938. Epub 2024 Jan 16. PMID: 38226681.

[20] Friedrich J, Rupp M, Feng YS, Sudeck G. Occupational health literacy and work ability: a moderation analysis including interpersonal and organizational factors in healthy organizations. Front Public Health. 2024 Feb 7;12:1243138. doi: 10.3389/fpubh.2024.1243138. PMID: 38384890; PMCID: PMC10879437.

[21] Powers JM, LaRowe LR, Garey L, Zvolensky MJ, Ditre JW. Pain intensity, e-cigarette dependence, and cessation-related outcomes: The moderating role of pain-related anxiety. Addict Behav. 2020 Dec;111:106548. doi: 10.1016/j.addbeh.2020.106548. Epub 2020 Jul 11. PMID: 32745941; PMCID: PMC7484173.

[22] Atif M, Asghar S, Mushtaq I, Malik I, Amin A, Babar ZU, Scahill S. What drives inappropriate use of antibiotics? A mixed methods study from Bahawalpur, Pakistan. Infect Drug Resist. 2019 Mar 26;12:687–699. doi: 10.2147/IDR.S189114. PMID: 30988635; PMCID: PMC6440533.

23. World Health Organization. (2020). Adverse Childhood Experiences International Questionnaire (ACE-IQ). https://www.who.int/publications/m/item/adverse-childhood-experiences-international-questionnaire-(ace-iq)

[24] Lin L, Wang HH, Lu C, Chen W, Guo VY. Adverse Childhood Experiences and Subsequent Chronic Diseases Among Middle-aged or Older Adults in China and Associations With Demographic and Socioeconomic Characteristics. JAMA Netw Open. 2021 Oct 1;4(10):e2130143. doi: 10.1001/jamanetworkopen.2021.30143. Erratum in: JAMA Netw Open. 2022 Jun 1;5(6):e2220614. PMID: 34694390; PMCID: PMC8546496.

[25] Huen JMY, Yip PSF, Osman A, Leung ANM. The Suicidal Behaviors Questionnaire-Revised (SBQ-R) and its Chinese version (C-SBQ-R): Further validity testing using the culture, comprehension, and translation bias procedure. Psychol Assess. 2022 Jul;34(7):704–710. doi: 10.1037/pas0001134. Epub 2022 Apr 25. PMID: 35467908.

[26] Zhou J, Tang R, Wang X, Li X, Heianza Y, Qi L. Improvement of Social Isolation and Loneliness and Excess Mortality Risk in People With Obesity. JAMA Netw Open. 2024 Jan 2;7(1):e2352824. doi: 10.1001/jamanetworkopen.2023.52824. PMID: 38252435; PMCID: PMC10804268.

[27] Elovainio M, Komulainen K, Sipilä PN, Pulkki-Råback L, Cachón Alonso L, Pentti J, Nyberg ST, Suominen S, Vahtera J, Lipsanen J, Batty GD, Hakulinen C, Kivimäki M. Association of social isolation and loneliness with risk of incident hospital-treated infections: an analysis of data from the UK Biobank and Finnish Health and Social Support studies. Lancet Public Health. 2023 Feb;8(2):e109–e118. doi: 10.1016/S2468-2667(22)00253-5. Epub 2023 Jan 17. PMID: 36669514; PMCID: PMC9879771.

[28] Hui Gan GZ, Hill AM, Yeung P, Keesing S, Netto JA. Pet ownership and its influence on mental health in older adults. Aging Ment Health. 2020 Oct;24(10):1605–1612. doi: 10.1080/13607863.2019.1633620. Epub 2019 Jun 27. PMID: 31242754.

[29] Gee NR, Rodriguez KE, Fine AH, Trammell JP. Dogs Supporting Human Health and Well-Being: A Biopsychosocial Approach. Front Vet Sci. 2021 Mar 30;8:630465. doi: 10.3389/fvets.2021.630465. PMID: 33860004; PMCID: PMC8042315.

[30] Pennings G. Elective egg freezing and women’s emancipation. Reprod Biomed Online. 2021 Jun;42(6):1053–1055. doi: 10.1016/j.rbmo.2021.04.004. Epub 2021 Apr 20. PMID: 33931374.

[31] Rajabalee NBMH, Joseph A, Tapper CX. Global Geriatric Palliative Care. Clin Geriatr Med. 2023 Aug;39(3):465–473. doi: 10.1016/j.cger.2023.05.002. PMID: 37385697.

[32] Phan H, Ngu B, Hsu CS, Chen SC. The Life + Death Education Framework: Proposition of a ‘Universal’ Framework for Implementation. Omega (Westport). 2024 Oct 26:302228241295786. doi: 10.1177/00302228241295786. Epub ahead of print. PMID: 39460748.

[33] Weilnhammer V, Schmid J, Mittermeier I, Schreiber F, Jiang L, Pastuhovic V, Herr C, Heinze S. Extreme weather events in europe and their health consequences - A systematic review. Int J Hyg Environ Health. 2021 Apr;233:113688. doi: 10.1016/j.ijheh.2021.113688. Epub 2021 Jan 30. PMID: 33530011.

[34] Rizan C, Steinbach I, Nicholson R, Lillywhite R, Reed M, Bhutta MF. The Carbon Footprint of Surgical Operations: A Systematic Review. Ann Surg. 2020 Dec;272(6):986–995. doi: 10.1097/SLA.0000000000003951. PMID: 32516230.

[35] Luo N, Liu G, Li M, Guan H, Jin X, Rand-Hendriksen K. Estimating an EQ-5D-5L Value Set for China. Value Health. 2017 Apr;20(4):662–669. doi: 10.1016/j.jval.2016.11.016. Epub 2017 Feb 9. PMID: 28408009.

[36] Koo M, Norman C, Chang H.Psychometric Evaluation of a Chinese Version of the eHealth Literacy Scale (eHEALS) in School Age Children. The international electronic journal of health education. 2012 ;15, 29–36.

[37] Jiang F, Wu Y. 2023 Report on eHealth Literacy of Chinese Residents [M]. Shanghai: Shanghai Jiao Tong University Press, 2024.

[38] Crandall A, Weiss-Laxer NS, Broadbent E, Holmes EK, Magnusson BM, Okano L, Berge JM, Barnes MD, Hanson CL, Jones BL, Novilla LB. The Family Health Scale: Reliability and Validity of a Short- and Long-Form. Front Public Health. 2020 Nov 20;8:587125. doi: 10.3389/fpubh.2020.587125. PMID: 33330329; PMCID: PMC7717993.

[39] Wang F, Wu Y, Sun X, Wang D, Ming WK, Sun X, Wu Y. Reliability and validity of the Chinese version of a short form of the family health scale. BMC Prim Care. 2022 May 6;23(1):108. doi: 10.1186/s12875-022-01702-1. PMID: 35524178; PMCID: PMC9077878.

[40] Duong TV, Aringazina A, Kayupova G, Nurjanah, Pham TV, Pham KM, Truong TQ, Nguyen KT, Oo WM, Su TT, Majid HA, Sørensen K, Lin IF, Chang Y, Yang SH, Chang PWS. Development and Validation of a New Short-Form Health Literacy Instrument (HLS-SF12) for the General Public in Six Asian Countries. Health Lit Res Pract. 2019 Apr 10;3(2):e91–e102. doi: 10.3928/24748307-20190225-01. PMID: 31294310; PMCID: PMC6607763.

[41] Sun X, Lv K, Wang F, Ge P, Niu Y, Yu W, Sun X, Ming WK, He M, Wu Y. Validity and reliability of the Chinese version of the Health Literacy Scale Short-Form in the Chinese population. BMC Public Health. 2023 Feb 23;23(1):385. doi: 10.1186/s12889-023-15237-2. PMID: 36823591; PMCID: PMC9951431.

[42] Sun X, Chen K, Wu Y, Tang J, Wang F, Sun X, He M, Wu Y. Development of a short version of the Health Literacy Scale based on classical test theory and item response theory. Chinese General Practice, 2024 Aug, 27(23):2931–2940.

[43] Jokić-Begić N, Mikac U, Čuržik D, Čuržik D. The Development and Validation of the Short Cyberchondria Scale (SCS). Journal of Psychopathology and Behavioral Assessment, 2019 May,41:662–676. 10.1007/s10862-019-09744-z.

[44] Zhou J, Tang R, Wang X, Li X, Heianza Y, Qi L. Improvement of Social Isolation and Loneliness and Excess Mortality Risk in People With Obesity. JAMA Netw Open. 2024 Jan 2;7(1):e2352824. doi: 10.1001/jamanetworkopen.2023.52824. PMID: 38252435; PMCID: PMC10804268.

[45] Elovainio M, Komulainen K, Sipilä PN, Pulkki-Råback L, Cachón Alonso L, Pentti J, Nyberg ST, Suominen S, Vahtera J, Lipsanen J, Batty GD, Hakulinen C, Kivimäki M. Association of social isolation and loneliness with risk of incident hospital-treated infections: an analysis of data from the UK Biobank and Finnish Health and Social Support studies. Lancet Public Health. 2023 Feb;8(2):e109–e118. doi: 10.1016/S2468-2667(22)00253-5. Epub 2023 Jan 17. PMID: 36669514; PMCID: PMC9879771.

[46] Sudlow C, Gallacher J, Allen N, Beral V, Burton P, Danesh J, Downey P, Elliott P, Green J, Landray M, Liu B, Matthews P, Ong G, Pell J, Silman A, Young A, Sprosen T, Peakman T, Collins R. UK biobank: an open access resource for identifying the causes of a wide range of complex diseases of middle and old age. PLoS Med. 2015 Mar 31;12(3):e1001779. doi: 10.1371/journal.pmed.1001779. PMID: 25826379; PMCID: PMC4380465.

[47] Hughes ME, Waite LJ, Hawkley LC, Cacioppo JT. A Short Scale for Measuring Loneliness in Large Surveys: Results From Two Population-Based Studies. Res Aging. 2004;26(6):655–672. doi: 10.1177/0164027504268574. PMID: 18504506; PMCID: PMC2394670.

[48] Foster HME, Gill JMR, Mair FS, Celis-Morales CA, Jani BD, Nicholl BI, Lee D, O’Donnell CA. Social connection and mortality in UK Biobank: a prospective cohort analysis. BMC Med. 2023 Nov 10;21(1):384. doi: 10.1186/s12916-023-03055-7. PMID: 37946218; PMCID: PMC10637015.

[49] He X, Li C, Qian J, Cui H, Wu W. Reliability and validity of a generalized anxiety disorder scale in general hospital outpatients. Shanghai Archives of Psychiatry. 2010 Aug;22(4):200–203.

[50] Shah SJ, Oreper S, Jeon SY, Boscardin WJ, Fang MC, Covinsky KE. Social Frailty Index: Development and validation of an index of social attributes predictive of mortality in older adults. Proc Natl Acad Sci U S A. 2023 Feb 14;120(7):e2209414120. doi: 10.1073/pnas.2209414120. Epub 2023 Feb 7. PMID: 36749720; PMCID: PMC9963593.

[51] John O, Donahue E, Kentle R. The Big Five Inventory-Versions 4a and 54. Berkeley, CA: University of California, Berkeley, Institute of Personality and Social Research. 1991.

[52] Rammstedt B, John OP. Measuring personality in one minute or less: A 10-item short version of the Big Five Inventory in English and German. Journal of research in Personality 2007 Feb 41(1): 203–212. doi: 10.1016/j.jrp.2006.02.001.

[53] Wang Y, Yao L, Liu L, Yang X, Wu H, Wang J, Wang L. The mediating role of self-efficacy in the relationship between Big five personality and depressive symptoms among Chinese unemployed population: a cross-sectional study. BMC Psychiatry. 2014 Mar 3;14:61. doi: 10.1186/1471-244X-14-61. PMID: 24581332; PMCID: PMC3976156.

[54] Kroenke K, Spitzer RL, Williams JB. The PHQ-9: validity of a brief depression severity measure. J Gen Intern Med. 2001 Sep;16(9):606–13. doi: 10.1046/j.1525-1497.2001.016009606.x. PMID: 11556941; PMCID: PMC1495268.

[55] Wang W, Bian Q, Zhao Y, Li X, Wang W, Du J, Zhang G, Zhou Q, Zhao M. Reliability and validity of the Chinese version of the Patient Health Questionnaire (PHQ-9) in the general population. Gen Hosp Psychiatry. 2014 Sep-Oct;36(5):539–44. doi: 10.1016/j.genhosppsych.2014.05.021. Epub 2014 Jun 6. PMID: 25023953.

[56] Kroenke K, Spitzer RL, Williams JB. The Patient Health Questionnaire-2: validity of a two-item depression screener. Med Care. 2003 Nov;41(11):1284–92. doi: 10.1097/01.MLR.0000093487.78664.3C. PMID: 14583691.

[57] Spitzer RL, Kroenke K, Williams JB, Löwe B. A brief measure for assessing generalized anxiety disorder: the GAD-7. Arch Intern Med. 2006 May 22;166(10):1092–7. doi: 10.1001/archinte.166.10.1092. PMID: 16717171.

[58] Wang F, Wu Y, Wang S, Du Z, Wu Y. Development of an optimal short form of the GAD-7 scale with cross-cultural generalizability based on Riskslim. Gen Hosp Psychiatry. 2024 Mar-Apr;87:33–40. doi: 10.1016/j.genhosppsych.2024.01.010. Epub 2024 Jan 26. PMID: 38301522.

[59] Cohen S, Kamarck T, Mermelstein R. A global measure of perceived stress. J Health Soc Behav. 1983 Dec;24(4):385–96. PMID: 6668417.

[60] Huang F, Wang H, Wang Z, Zhang J, Du W, Su C, Jia X, Ouyang Y, Wang Y, Li L, Jiang H, Zhang B. Psychometric properties of the perceived stress scale in a community sample of Chinese. BMC Psychiatry. 2020 Mar 20;20(1):130. doi: 10.1186/s12888-020-02520-4. Erratum in: BMC Psychiatry. 2020 May 25;20(1):260. PMID: 32197589; PMCID: PMC7082906.

[61] Blumenthal JA, Burg MM, Barefoot J, Williams RB, Haney T, Zimet G. Social support, type A behavior, and coronary artery disease. Psychosom Med. 1987 Jul-Aug;49(4):331-40. doi: 10.1097/00006842-198707000-00002. PMID: 3615762.

[62] Jiang Q. Perceived social support scale. Chinese Journal of Behavioral Medical Science. 2001; 10(10): 41–43.

[63] Wu Y, Tang J, Du Z, Chen K, Zhang X, Wang F, Sun X, Sun X, Wu Y. Development of a Short Version of the Perceived Social Support Scale: Based on Classical Test Theory and Item Response Theory. The 25th National Academic Conference of Psychology. 2023 Oct;3.doi:10.26914/c.cnkihy.2023.057053.

[64] Schwarzer R, Jerusalem M. Weinman J, Wright S, Johnston M. Generalized Self-Efficacy Scale. Measures in Health Psychology: A User’s Portfolio. Causal and control beliefs Windsor. 1995 Jan;35(37): 82–003.

[65] Chen G, Gully S, Eden D. Validation of a New General Self-Efficacy Scale. Organizational Research Methods. 2001 Jan;4(1): 62–83. doi:10.1177/109442810141004

[66] Feng X, Chen X. Reliability and Validity of New General Self - efficacy Scale(NGSSE). Journal of Mudanjiang Normal University (Philosophy and Social Sciences Edition). 2012 Aug;4(170):127–133. doi: 10.13815/j.cnki.jmtc(pss).2012.04.042

[67] Chiu CY, Hong YY, Dweck CS. Lay dispositionism and implicit theories of personality. J Pers Soc Psychol. 1997 Jul;73(1):19–30. doi: 10.1037//0022-3514.73.1.19. PMID: 9216077.

[68] Bunda K, Busseri MA. Lay theories of health, self-rated health, and health behavior intentions. J Health Psychol. 2019 Jun;24(7):979–988. doi: 10.1177/1359105316689143. Epub 2017 Jan 30. PMID: 28810398.

[69] Back M, Küfner A, Dufner M, Gerlach T, Rauthmann J, Denissen J. Narcissistic admiration and rivalry: Disentangling the bright and dark sides of narcissism. Journal of Personality and Social Psychology, 2013 Dec;105(6), 1013–1037. 10.1037/a0034431.

[70] Leckelt M, Wetzel E, Gerlach TM, Ackerman RA, Miller JD, Chopik WJ, Penke L, Geukes K, Küfner ACP, Hutteman R, Richter D, Renner KH, Allroggen M, Brecheen C, Campbell WK, Grossmann I, Back MD. Validation of the Narcissistic Admiration and Rivalry Questionnaire Short Scale (NARQ-S) in convenience and representative samples. Psychol Assess. 2018 Jan;30(1):86–96. doi: 10.1037/pas0000433. Epub 2017 Mar 2.

[71] Ustun B, Adler LA, Rudin C, Faraone SV, Spencer TJ, Berglund P, Gruber MJ, Kessler RC. The World Health Organization Adult Attention-Deficit/Hyperactivity Disorder Self-Report Screening Scale for DSM-5. JAMA Psychiatry. 2017 May 1;74(5):520–527. doi: 10.1001/jamapsychiatry.2017.0298. Erratum in: JAMA Psychiatry. 2017 Dec 1;74(12):1279. Erratum in: JAMA Psychiatry. 2019 Nov 1;76(11):1213. PMID: 28384801; PMCID: PMC5470397.

[72] Borritz M, Bültmann U, Rugulies R, Christensen K, Villadsen E, Kristensen T. Psychosocial Work Characteristics as Predictors for Burnout: Findings From 3-Year Follow Up of the PUMA Study. Journal of Occupational and Environmental Medicine. 2005 Oct;47(10), 1015–1025. 10.1097/01.jom.0000175155.50789.98.

[73] Borritz M, Bültmann U, Rugulies R, Christensen K, Villadsen E, Kristensen T. Psychosocial Work Characteristics as Predictors for Burnout: Findings From 3-Year Follow Up of the PUMA Study. Journal of Occupational and Environmental Medicine, 2005 Oct;47(10), 1015–1025. http://www.jstor.org/stable/44998861.

[74] Wang F, Song H, Meng X, Wang T, Zhang Q, Yu Z, Fan S, Wu Y. Development and validation of the long and short forms of the rest intolerance scale for college students. Personality and Individual Differences. 2025 Feb;233:112869. 10.1016/j.paid.2024.112869

[75] Somer E, Lehrfeld J, Bigelsen J, Jopp DS. Development and validation of the Maladaptive Daydreaming Scale (MDS). Conscious Cogn. 2016 Jan;39:77–91. doi: 10.1016/j.concog.2015.12.001. Epub 2015 Dec 17. PMID: 26707384.

[76] Pietkiewicz IJ, Hełka AM, Barłóg M, Tomalski R. Validity and reliability of the Polish Maladaptive Daydreaming Scale (PMDS-16) and its short form (PMDS-5). Clin Psychol Psychother. 2023 Jul-Aug;30(4):882–897. doi: 10.1002/cpp.2844. Epub 2023 Mar 10. PMID: 36809856.

[77] Pietkiewicz IJ, Hełka AM, Barłóg M, Tomalski R. Validity and reliability of the Polish Maladaptive Daydreaming Scale (PMDS-16) and its short form (PMDS-5). Clin Psychol Psychother. 2023 Jul-Aug;30(4):882–897. doi: 10.1002/cpp.2844. Epub 2023 Mar 10. PMID: 36809856.

[78] Bányai F, Zsila Á, Király O, Maraz A, Elekes Z, Griffiths MD, Andreassen CS, Demetrovics Z. Problematic Social Media Use: Results from a Large-Scale Nationally Representative Adolescent Sample. PLoS One. 2017 Jan 9;12(1):e0169839. doi: 10.1371/journal.pone.0169839. PMID: 28068404; PMCID: PMC5222338.

[79] Leung H, Pakpour AH, Strong C, Lin YC, Tsai MC, Griffiths MD, Lin CY, Chen IH. Measurement invariance across young adults from Hong Kong and Taiwan among three internet-related addiction scales: Bergen Social Media Addiction Scale (BSMAS), Smartphone Application-Based Addiction Scale (SABAS), and Internet Gaming Disorder Scale-Short Form (IGDS-SF9) (Study Part A). Addict Behav. 2020 Feb;101:105969. doi: 10.1016/j.addbeh.2019.04.027. Epub 2019 Apr 27. PMID: 31078344.

[80] Crumbaugh, J. Manual of instruction for the Purpose-in-Life Test. Psychometric affiliates. 1969.

[81] Zhang D, Chan DC, Niu L, Liu H, Zou D, Chan AT, Gao TT, Zhong B, Sit RW, Wong SY. Meaning and its association with happiness, health and healthcare utilization: A cross-sectional study. J Affect Disord. 2018 Feb;227:795–802. doi: 10.1016/j.jad.2017.11.082. Epub 2017 Nov 16. PMID: 29689694.

[82] Nosikov A, Gudex. Development of a common instrument for physical activity. EUROHIS: develo** common instruments for health surveys. 2003; 79.

[83] Hu B, Lin L, Yuan Z, Yang Y, Lv M, Ye W, Yu S, L J. A validity study of three physical activity questionnaires. Modern Preventive Medicine. 2013 Nov;40(16):3061–3065.

[84] Yao Y, Jia Y, Wen Y, Cheng B, Cheng S, Liu L, Yang X, Meng P, Chen Y, Li C, Zhang J, Zhang Z, Pan C, Zhang H, Wu C, Wang X, Ning Y, Wang S, Zhang F. Genome-Wide Association Study and Genetic Correlation Scan Provide Insights into Its Genetic Architecture of Sleep Health Score in the UK Biobank Cohort. Nat Sci Sleep. 2022 Jan 6;14:1–12. doi: 10.2147/NSS.S326818. PMID: 35023977; PMCID: PMC8747788.

[85] Fan M, Sun D, Zhou T, Heianza Y, Lv J, Li L, Qi L. Sleep patterns, genetic susceptibility, and incident cardiovascular disease: a prospective study of 385-292 UK biobank participants. Eur Heart J. 2020 Mar 14;41(11):1182–1189. doi: 10.1093/eurheartj/ehz849. PMID: 31848595; PMCID: PMC7071844.

[86] Olson DH, Sprenkle DH, Russell CS. Circumplex model of marital and family system: I. Cohesion and adaptability dimensions, family types, and clinical applications. Fam Process. 1979 Mar;18(1):3–28. doi: 10.1111/j.1545-5300.1979.00003.x. PMID: 437067.

[87] Guo N, Ho HCY, Wang MP, Lai AY, Luk TT, Viswanath K, Chan SS, Lam TH. Factor Structure and Psychometric Properties of the Family Communication Scale in the Chinese Population. Front Psychol. 2021 Nov 12;12:736514. doi: 10.3389/fpsyg.2021.736514. PMID: 34867617; PMCID: PMC8632692.

[88] Kliemann N, Beeken RJ, Wardle J, Johnson F. Development and validation of the Self-Regulation of Eating Behaviour Questionnaire for adults. Int J Behav Nutr Phys Act. 2016 Aug 2;13:87. doi: 10.1186/s12966-016-0414-6. PMID: 27484457; PMCID: PMC4969721.

[89] Bover MT, Foulds J, Steinberg MB, Richardson D, Marcella SW. Waking at night to smoke as a marker for tobacco dependence: patient characteristics and relationship to treatment outcome. Int J Clin Pract. 2008 Feb;62(2):182–90. doi: 10.1111/j.1742-1241.2007.01653.x. PMID: 18199277.

[90] Lin L, Wang HH, Lu C, Chen W, Guo VY. Adverse Childhood Experiences and Subsequent Chronic Diseases Among Middle-aged or Older Adults in China and Associations With Demographic and Socioeconomic Characteristics. JAMA Netw Open. 2021 Oct 1;4(10):e2130143. doi: 10.1001/jamanetworkopen.2021.30143. Erratum in: JAMA Netw Open. 2022 Jun 1;5(6):e2220614. PMID: 34694390; PMCID: PMC8546496.

[91] Fujiwara T. Impact of adverse childhood experience on physical and mental health: A life-course epidemiology perspective. Psychiatry Clin Neurosci. 2022 Nov;76(11):544–551. doi: 10.1111/pcn.13464. Epub 2022 Sep 24. PMID: 36002401.

[92] Mittal C, Griskevicius V, Simpson JA, Sung S, Young ES. Cognitive adaptations to stressful environments: When childhood adversity enhances adult executive function. J Pers Soc Psychol. 2015 Oct;109(4):604–621. doi: 10.1037/pspi0000028. PMID: 26414842.

[93] van Minde MRC, de Kroon MLA, Sijpkens MK, Raat H, Steegers EAP, Bertens LCM. Associations between Socio-Economic Status and Unfavorable Social Indicators of Child Wellbeing; a Neighbourhood Level Data Design. Int J Environ Res Public Health. 2021 Dec 1;18(23):12661. doi: 10.3390/ijerph182312661. PMID: 34886386; PMCID: PMC8657207.

[94] Gur A, Rimmerman A. Social involvement, socio-economic status and subjective well-being of parents of offspring with intellectual and developmental disabilities. J Intellect Disabil Res. 2021 Sep;65(9):870–877. doi: 10.1111/jir.12841. Epub 2021 May 6. PMID: 33955605.

[95] Ministry of Education of the People’s Republic of China. (2007). The Ministry of Education’s Opinions on Strengthening and Improving Art Education Activities in Primary and Secondary Schools. http://www.moe.gov.cn/srcsite/A17/moe_794/moe_795/200705/t20070530_80592.html

